## Supplementary Figures and Text for "A framework for conducting time-varying genome-wide association studies: An application to body mass index across childhood in six multiethnic cohorts"

### Supplementary Figure 1: Flow diagram of analysis pipeline

The flow diagram illustrates the step by step procedure used to implement, test, assess and finalize non-linear BMI curve modeling ready for GWAS. Future analysts may consult this flow diagram and read further information in the Supplementary text about the outputs from the *EGGLA* pipeline.

*ALSPAC Avon Longitudinal Study of Parents and Children; CHOP-EA Children's Hospital of Philadelphia European subset; CHOP-AA Children's Hospital of Philadelphia African American subset; NFBC1966 Northern Finland Birth Cohort 1966; NFBC1986 Northern Finland Birth Cohort 1986; LMM linear mixed model; GWAS genome-wide association study; BMI body mass index; glm generalized linear model; PCs principal components; QC quality control. *Here model refinement is specific to the present BMI growth analysis. For other growth methods, model refinement will be specific to the chosen model and knot refinements for spline models may not be appropriate.*

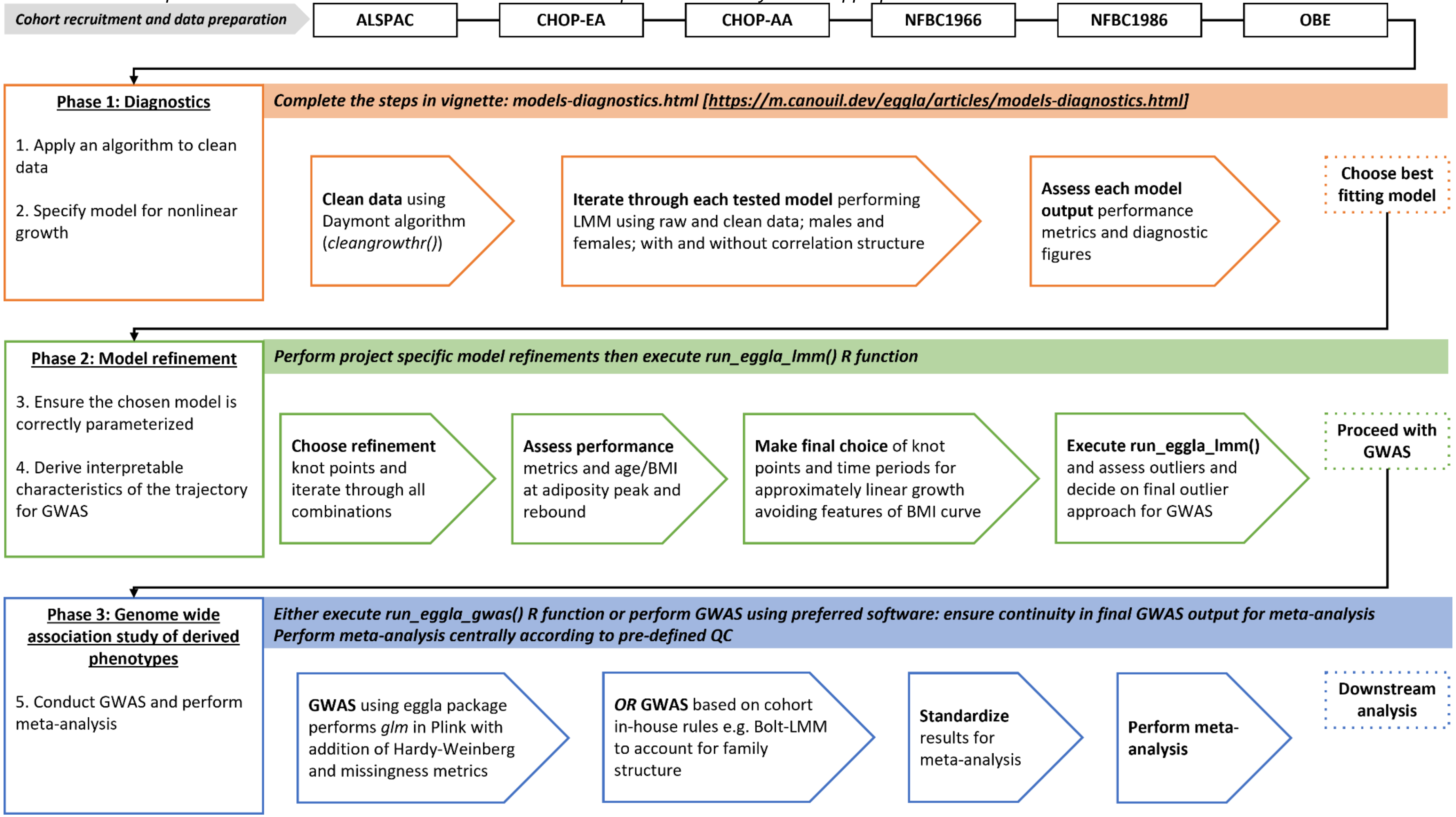

### Supplementary Figure 2: Refinement of knot points in the cubic spline with cubic slope random effects model

The cubic spline with cubic slope random effects models were selected for refinement. Models iterated pairwise through increments of knot 1 (1, 1.5 and 2 years) and knot 2 (6, 7, and 8 years), with knot 3 being held constant at 12 years. The figure displays the performance metrics AIC, conditional $R^2$, and RMSE as a function of average age at adiposity peak and rebound. The knot point combinations are differentially coloured for knot 1 (yellow for 1 year, blue for 1.5 years, and green for 2 years) and has a differential marker symbol to denote knot 2 (circle for 6 years, square for 7 years, and triangle for 8 years). Marker colour indicates males (blue) and females (pink). Figures are given for ALSPAC [A], CHOP-EA [B], CHOP-AA [C], NFBC1966 [D], NFBC1986 [E], and OBE [F].

*ALSPAC Avon Longitudinal Study of Parents and Children; CHOP-EA Children's Hospital of Philadelphia European subset; CHOP-AA Children's Hospital of Philadelphia African American subset; NFBC1966 Northern Finland Birth Cohort 1966; NFBC1986 Northern Finland Birth Cohort 1986; AIC Akaike information criterion ; RMSE root mean square error.*

#### A: ALSPAC

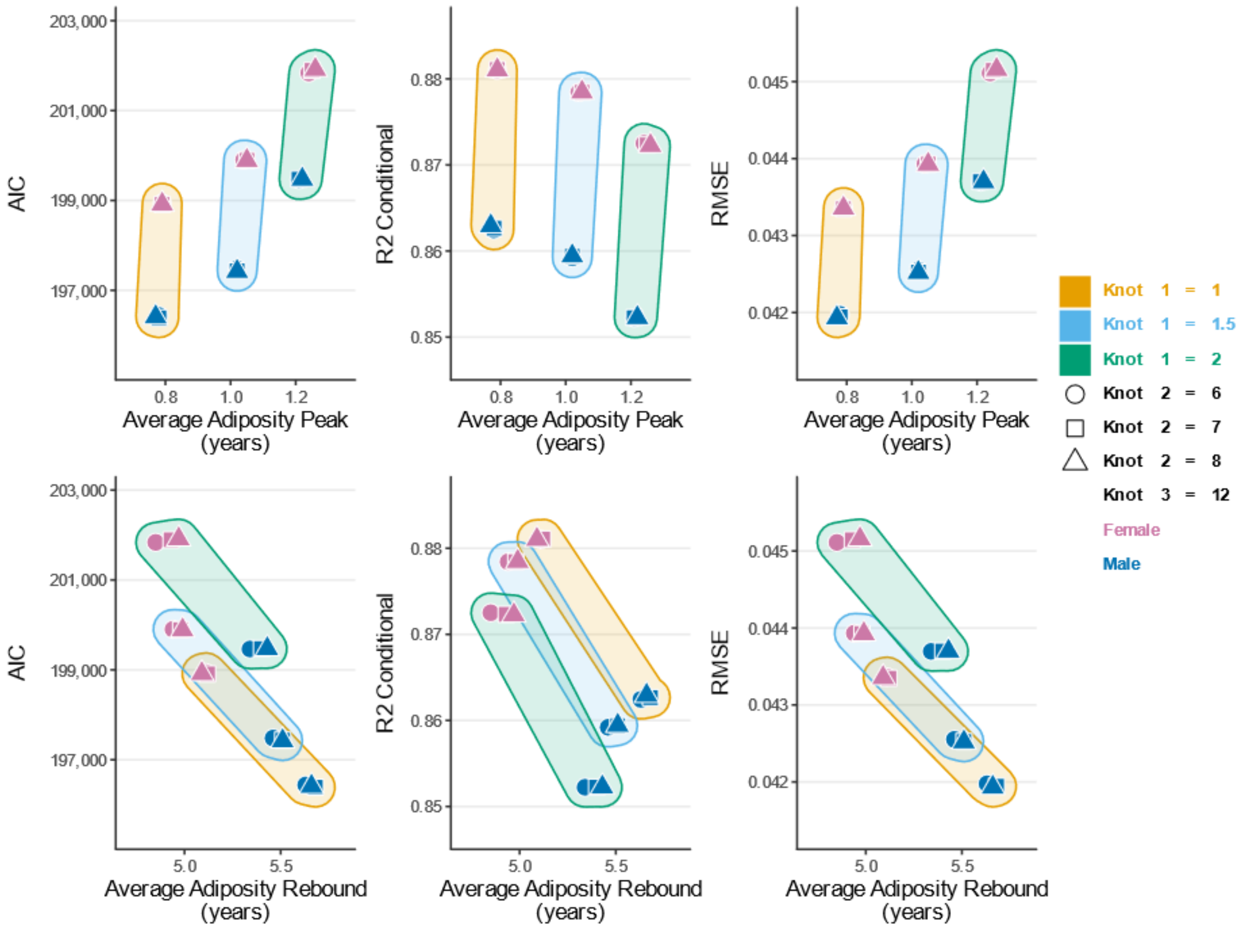

B: CHOP-EA

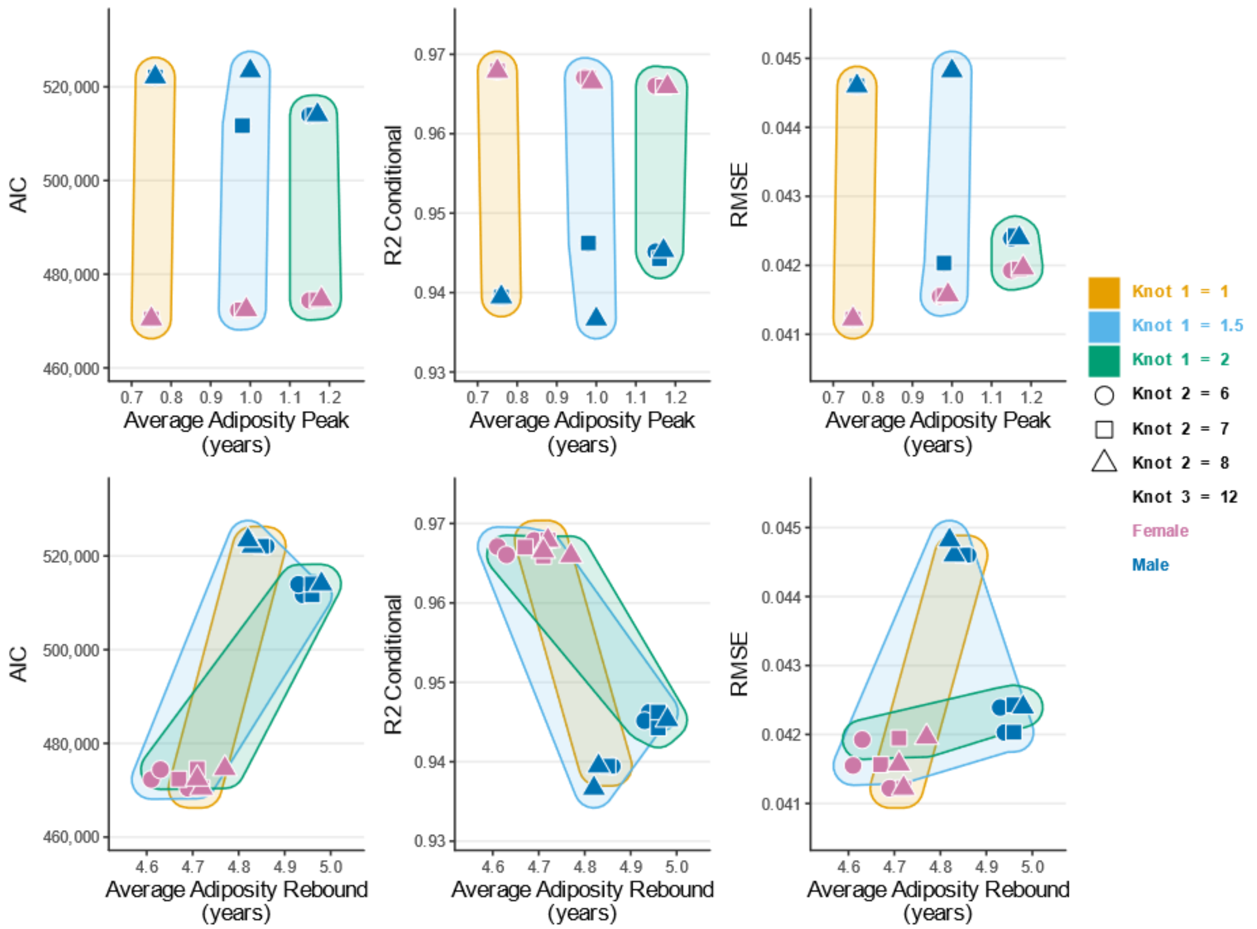

AIC
520,000
500,000
480,000
460,000
R2 Conditional
0.97
0.96
0.95
0.94
0.93
RMSE
0.045
0.044
0.043
0.042
0.041
0.7 0.8 0.9 1.0 1.1 1.2
Average Adiposity Peak (years)
4.6 4.7 4.8 4.9 5.0
Average Adiposity Rebound (years)
Knot 1 = 1
Knot 1 = 1.5
Knot 1 = 2
Knot 2 = 6
Knot 2 = 7
Knot 2 = 8
Knot 3 = 12
Female
Male

C: CHOP-AA

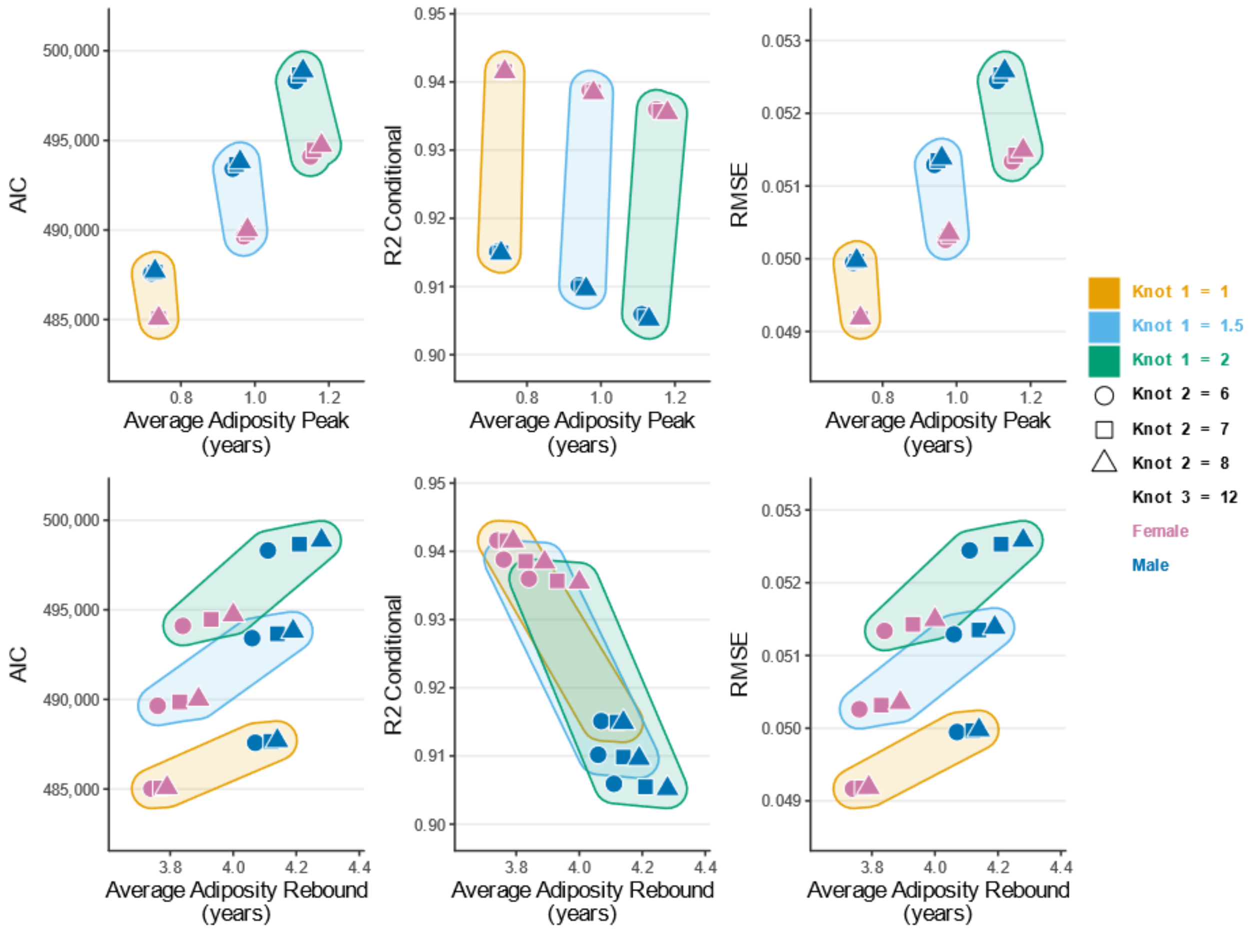

AIC
500,000
495,000
490,000
485,000
0.8
1.0
1.2
Average Adiposity Peak (years)
R2 Conditional
0.95
0.94
0.93
0.92
0.91
0.90
RMSE
0.053
0.052
0.051
0.050
0.049
Average Adiposity Rebound (years)
3.8
4.0
4.2
4.4
Knot 1 = 1
Knot 1 = 1.5
Knot 1 = 2
Knot 2 = 6
Knot 2 = 7
Knot 2 = 8
Knot 3 = 12
Female
Male

D: NFBC1966

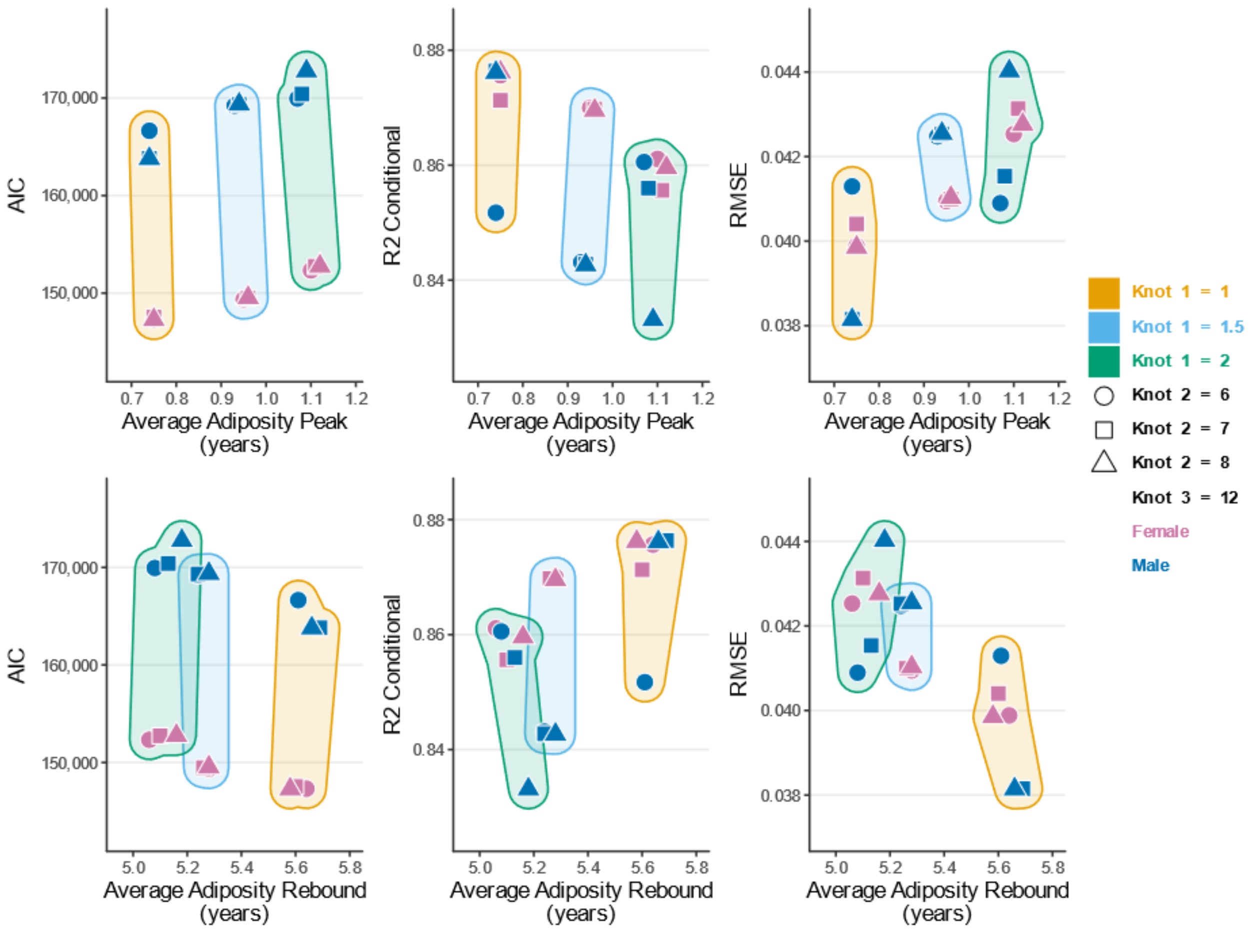

AIC
170,000
160,000
150,000
R2 Conditional
0.88
0.86
0.84
RMSE
0.044
0.042
0.040
0.038
0.7 0.8 0.9 1.0 1.1 1.2
Average Adiposity Peak (years)
5.0 5.2 5.4 5.6 5.8
Average Adiposity Rebound (years)
Knot 1 = 1
Knot 1 = 1.5
Knot 1 = 2
Knot 2 = 6
Knot 2 = 7
Knot 2 = 8
Knot 3 = 12
Female
Male

E: NFBC1986

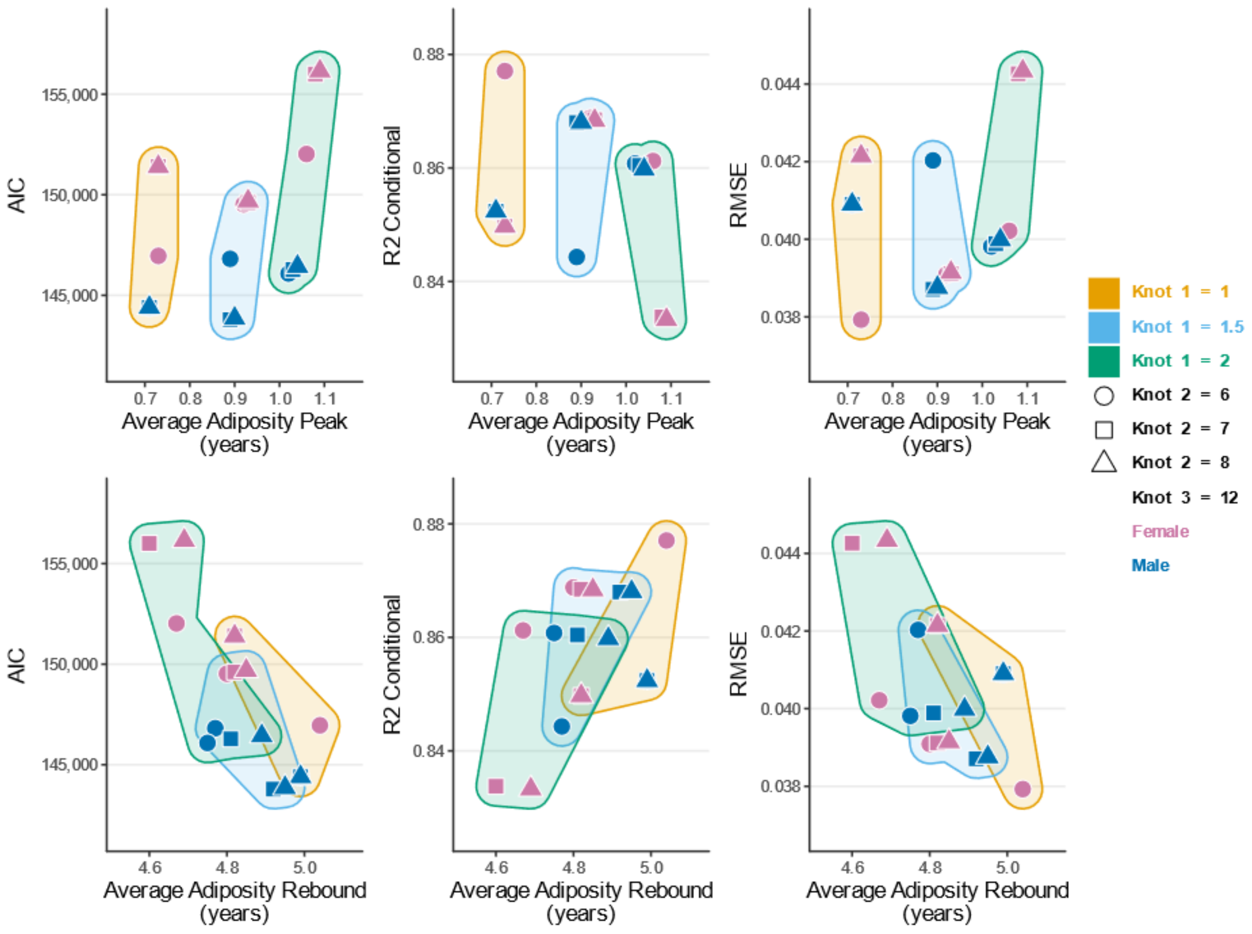

AIC
R2 Conditional
RMSE
Average Adiposity Peak (years)
Average Adiposity Rebound (years)
Knot 1 = 1
Knot 1 = 1.5
Knot 1 = 2
Knot 2 = 6
Knot 2 = 7
Knot 2 = 8
Knot 3 = 12
Female
Male

F: OBE

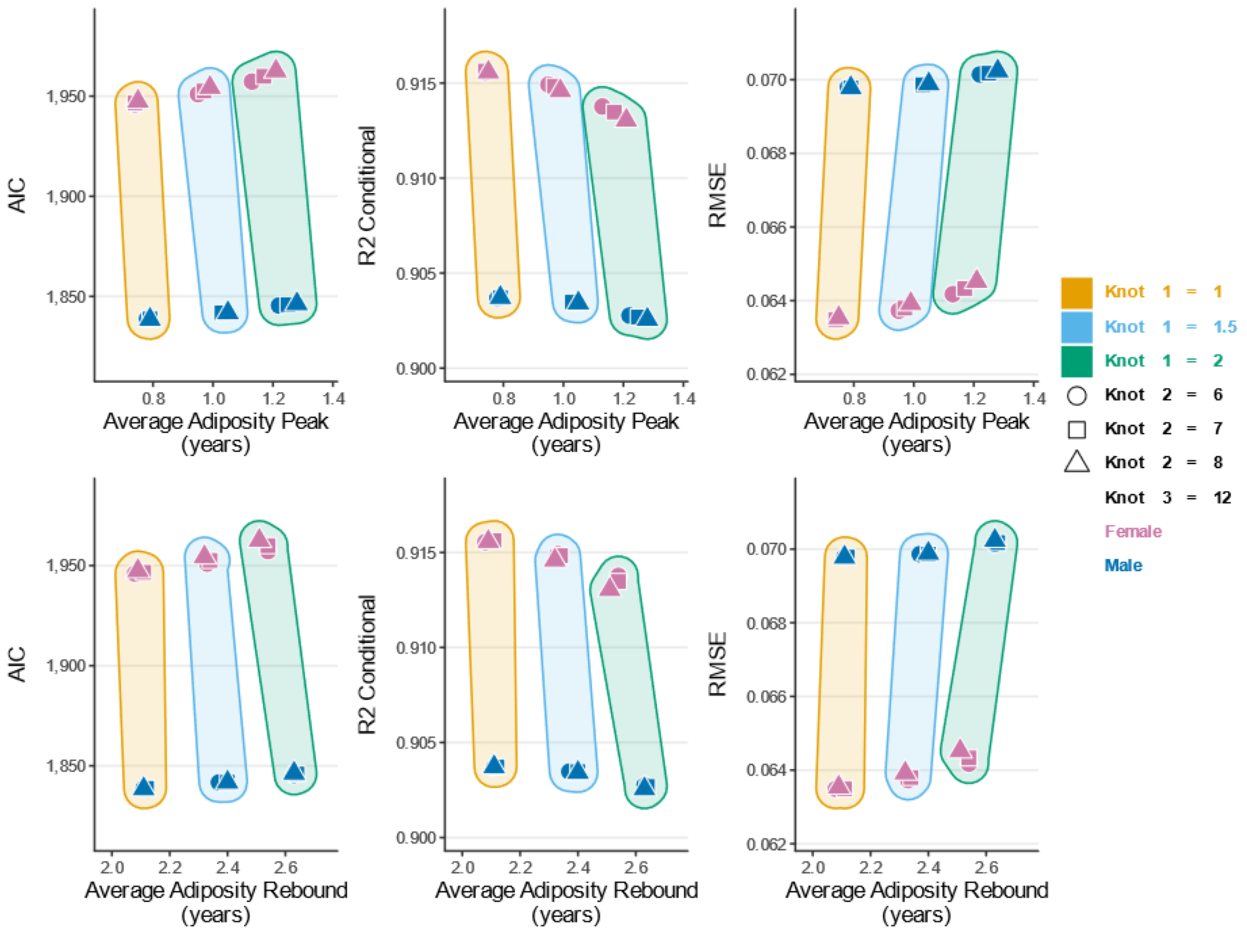
AIC
R2 Conditional
RMSE
Average Adiposity Peak (years)
Average Adiposity Rebound (years)
Knot 1 = 1
Knot 1 = 1.5
Knot 1 = 2
Knot 2 = 6
Knot 2 = 7
Knot 2 = 8
Knot 3 = 12
Female
Male

### Supplementary Figure 3: Residual plots for the chosen model

Assessment of model fit for our final chosen model (cubic spline with cubic slope random effects). Each plot depicts: A) the fitted BMI values vs. observed B) the fitted BMI values vs. marginal residuals C) the fitted age values vs. marginal residuals D) autocorrelation function (ACF) vs. lag of normalized residuals E) theoretical vs. sample quantiles and F) theoretical vs. residual quantiles. 1) ALSPAC; 2) CHOP-EA; 3) CHOP-AA 4) NFBC1966; 5) NFBC1986; 6) OBE.

*ALSPAC Avon Longitudinal Study of Parents and Children; CHOP-EA Children's Hospital of Philadelphia European subset; CHOP-AA Children's Hospital of Philadelphia African American subset; NFBC1966 Northern Finland Birth Cohort 1966; NFBC1986 Northern Finland Birth Cohort 1986*

#### 1: ALSPAC

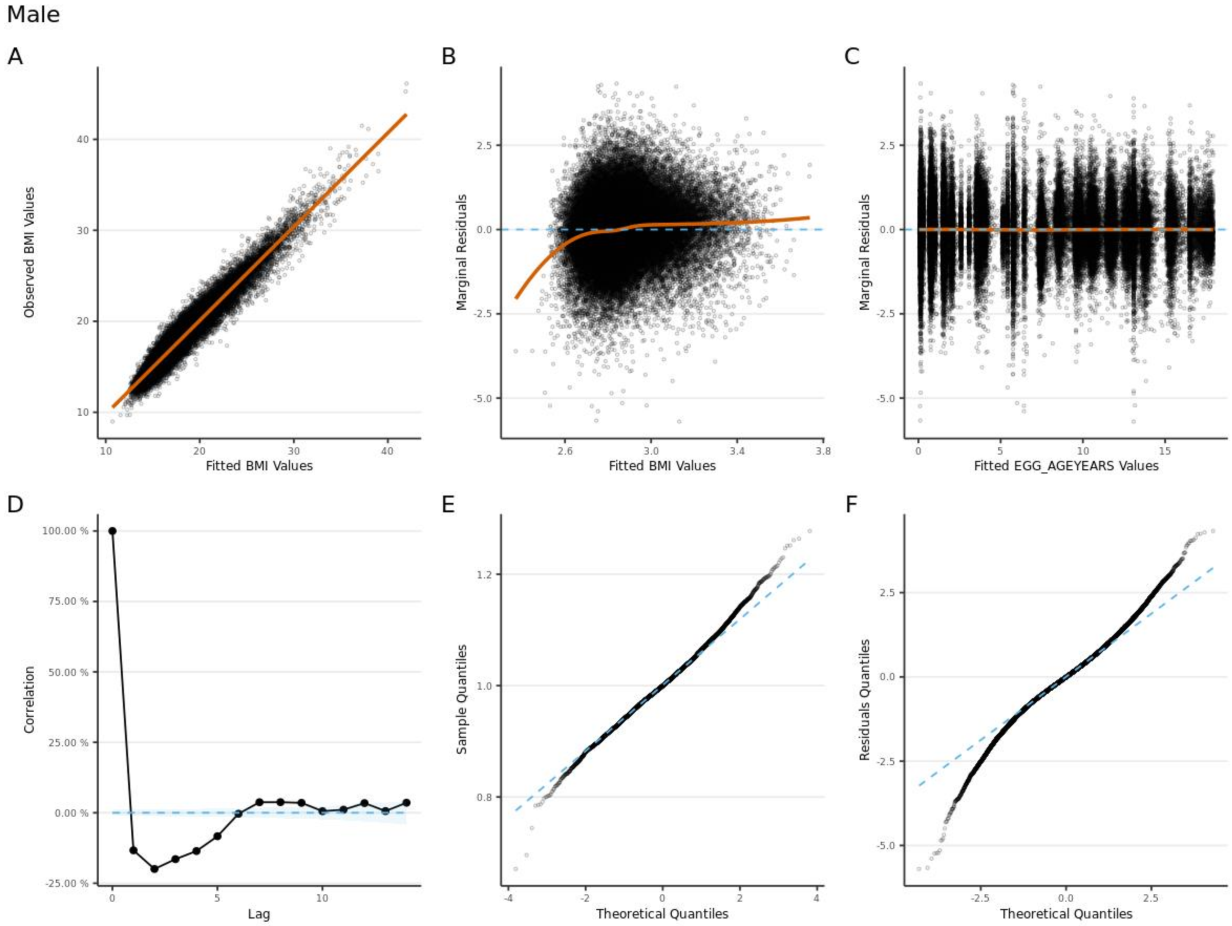

Female

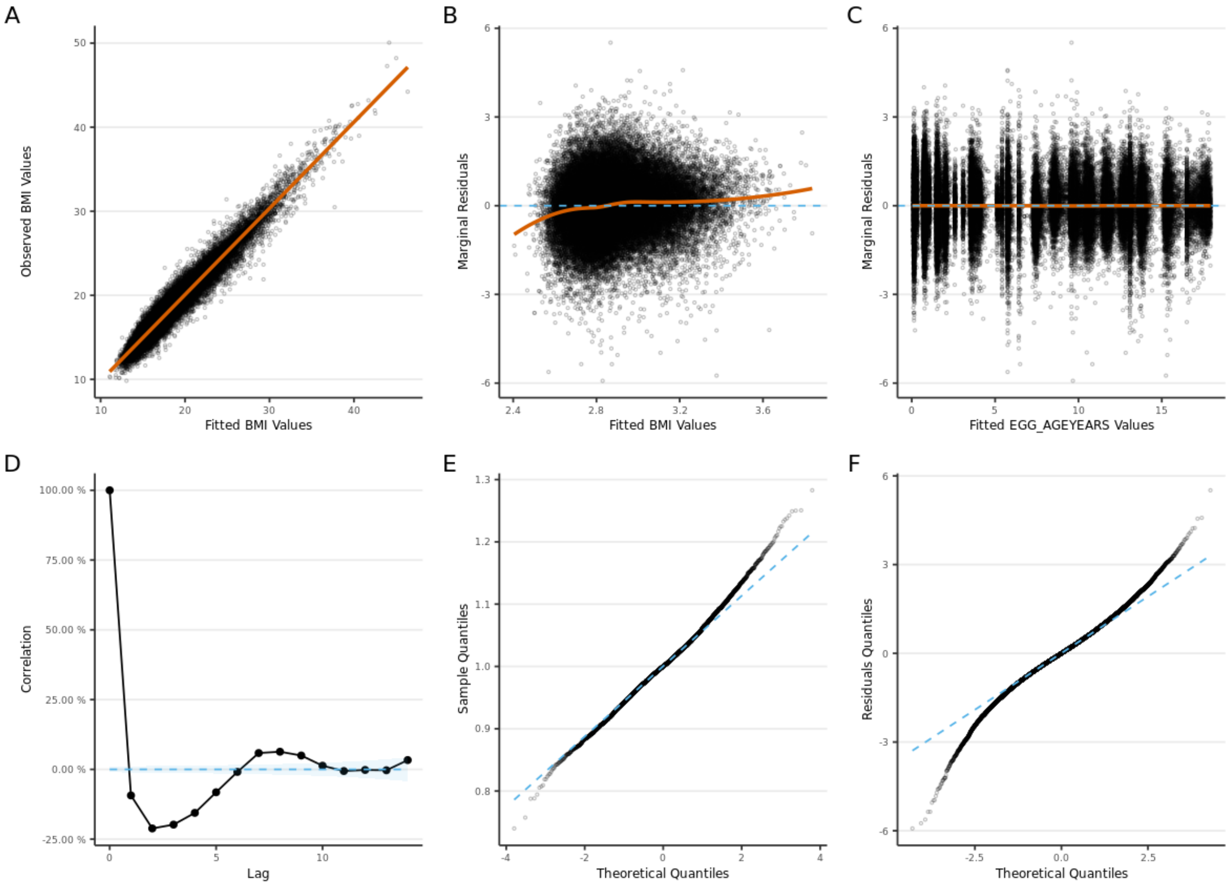
A
B
C
D
E
F
Observed BMI Values
Fitted BMI Values
Marginal Residuals
Fitted BMI Values
Marginal Residuals
Fitted EGG_AGEYEARS Values
Correlation
Lag
Sample Quantiles
Theoretical Quantiles
Residuals Quantiles
Theoretical Quantiles

### 2: CHOP-EA

#### Male

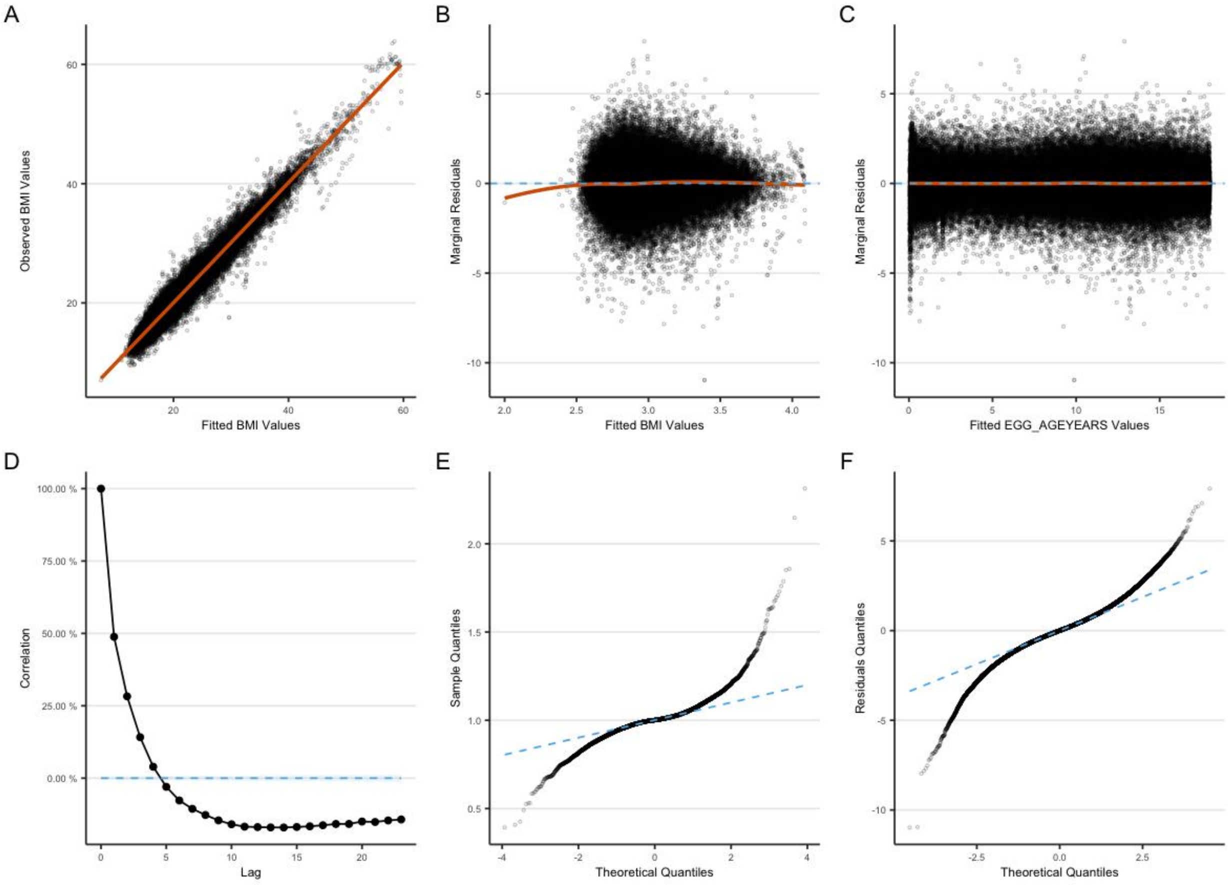

Female

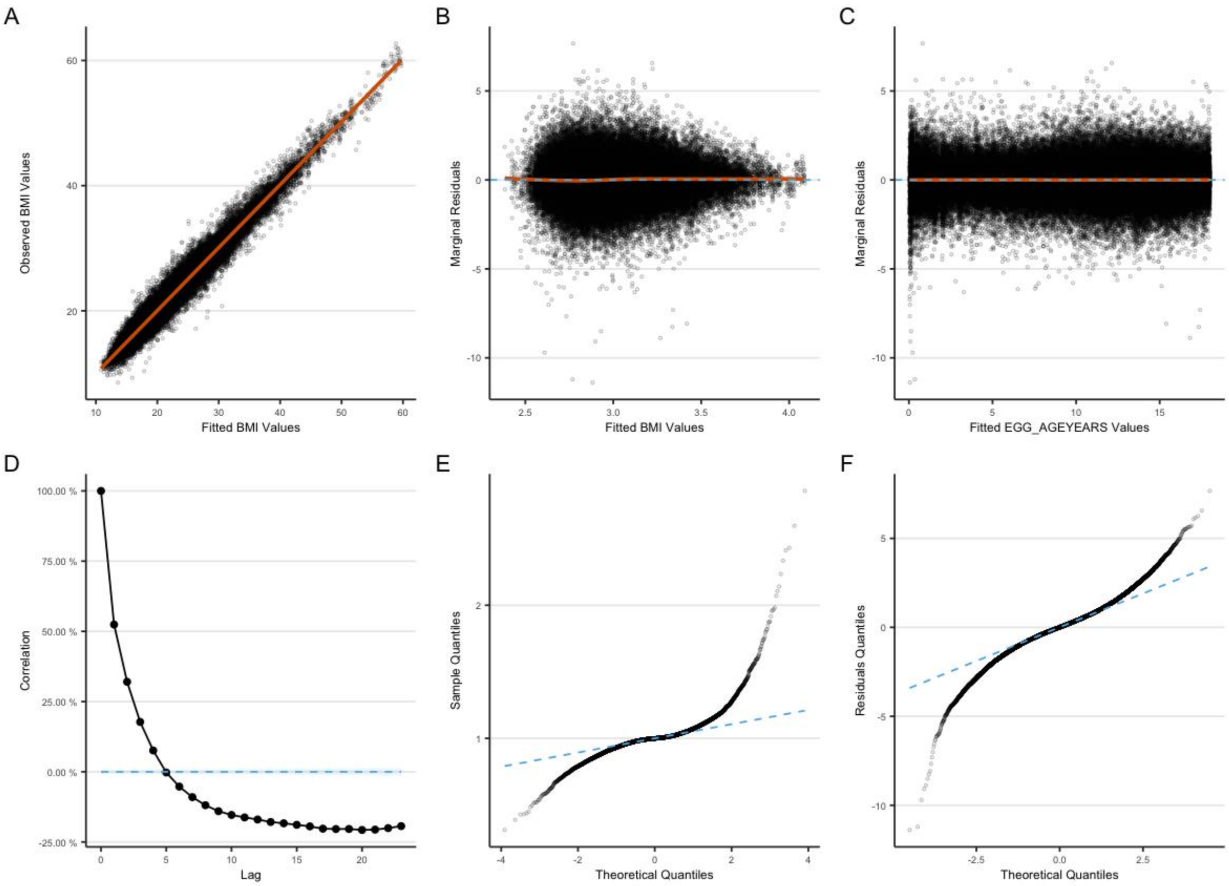

A
Observed BMI Values
Fitted BMI Values
B
Marginal Residuals
Fitted BMI Values
C
Marginal Residuals
Fitted EGG_AGEYEARS Values
D
Correlation
Lag
E
Sample Quantiles
Theoretical Quantiles
F
Residuals Quantiles
Theoretical Quantiles

### 3: CHOP-AA

#### Male

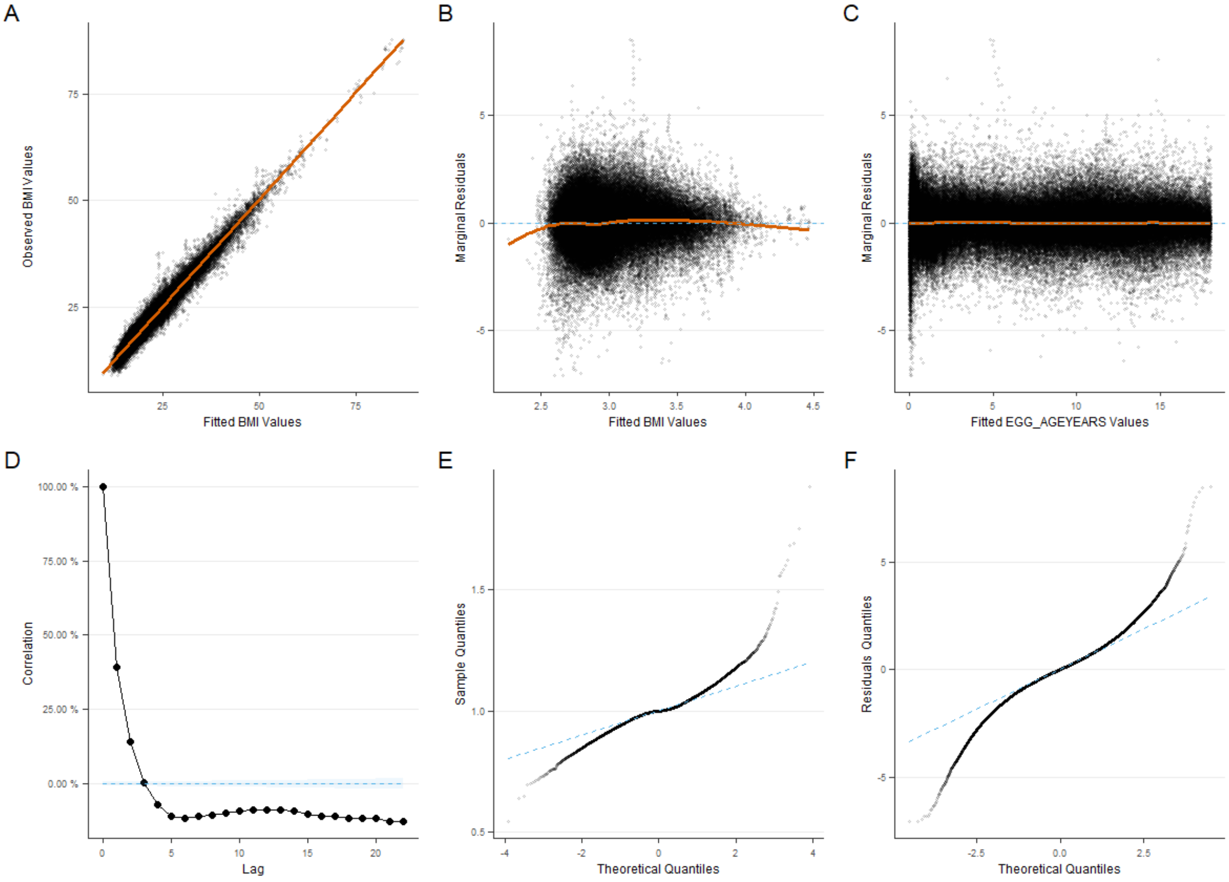

Female

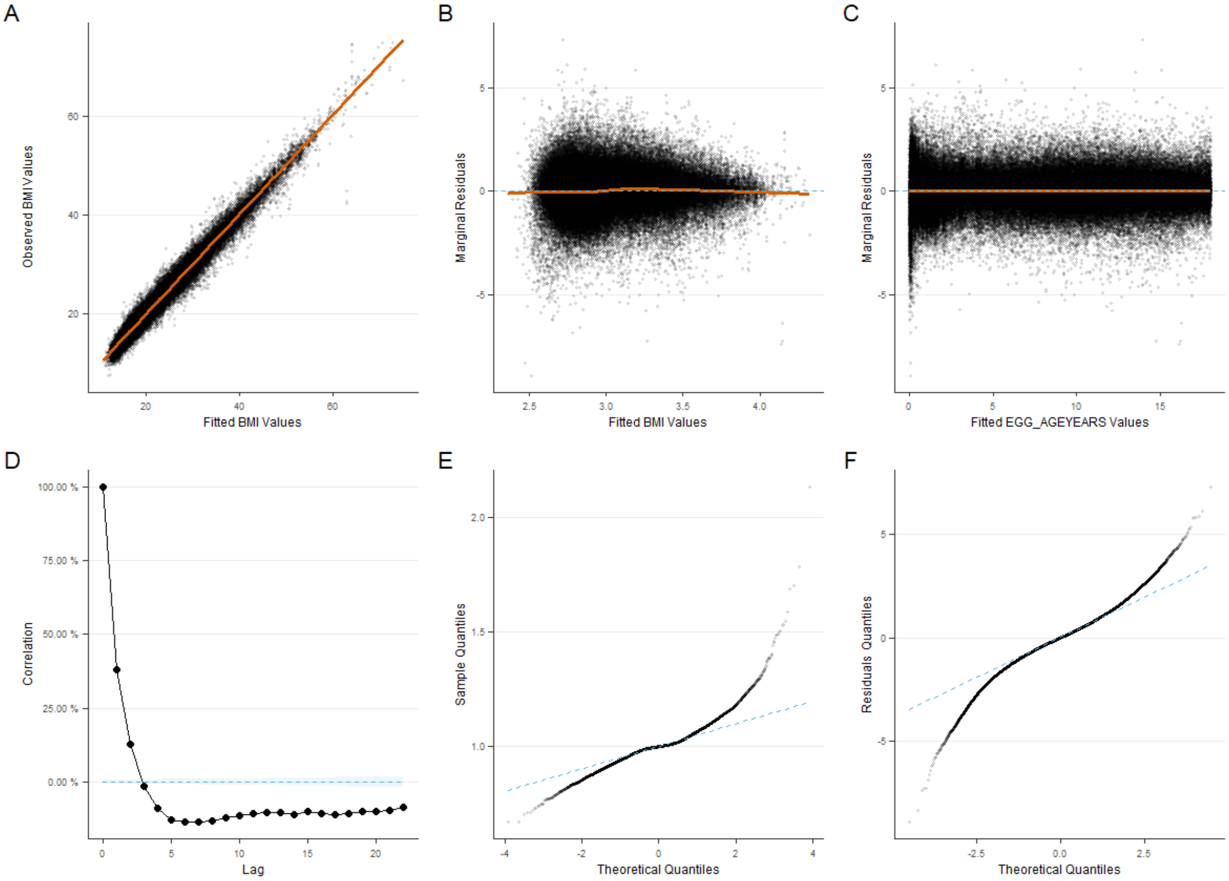
A
B
C
D
E
F
Observed BMI Values
Fitted BMI Values
Marginal Residuals
Fitted BMI Values
Marginal Residuals
Fitted EGG_AGEYEARS Values
Correlation
Lag
Sample Quantiles
Theoretical Quantiles
Residuals Quantiles
Theoretical Quantiles

4: NFBC1966

Male

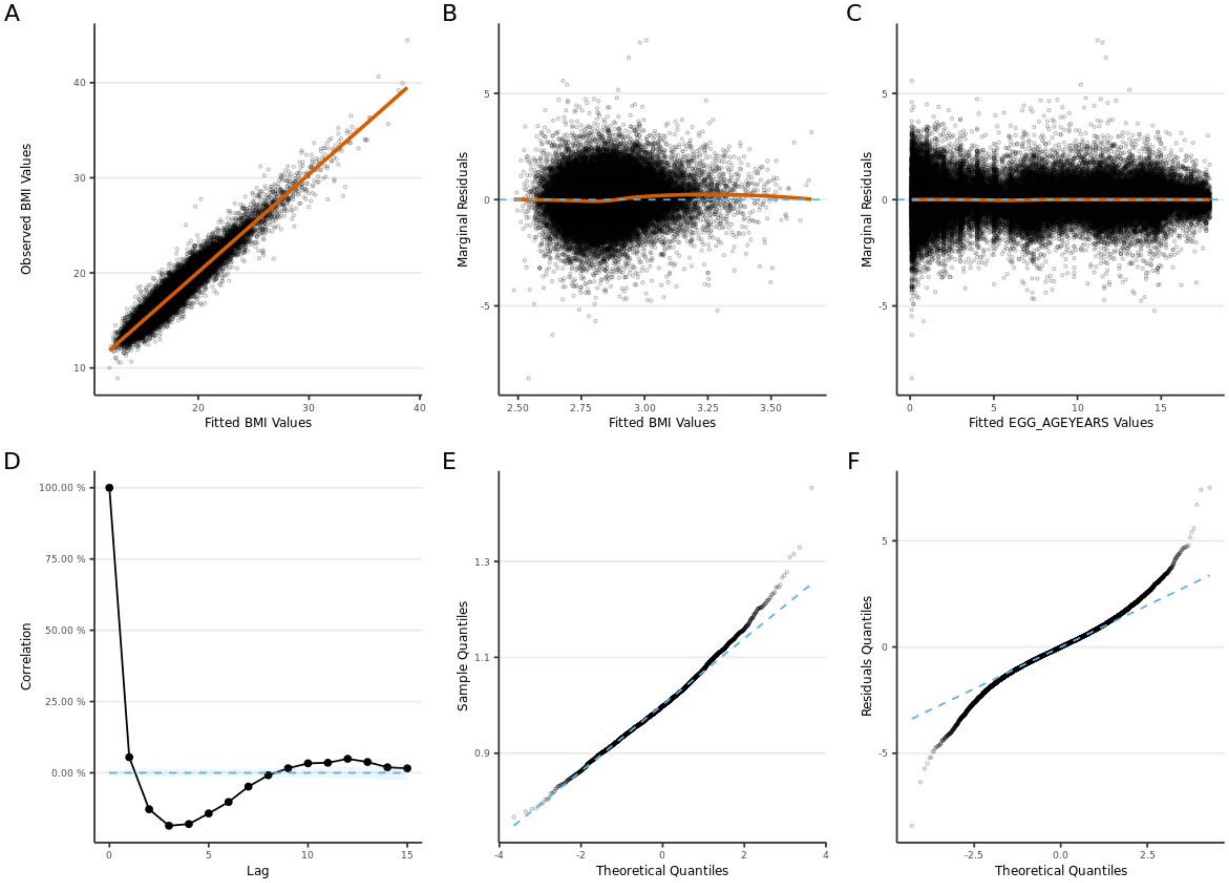

Female

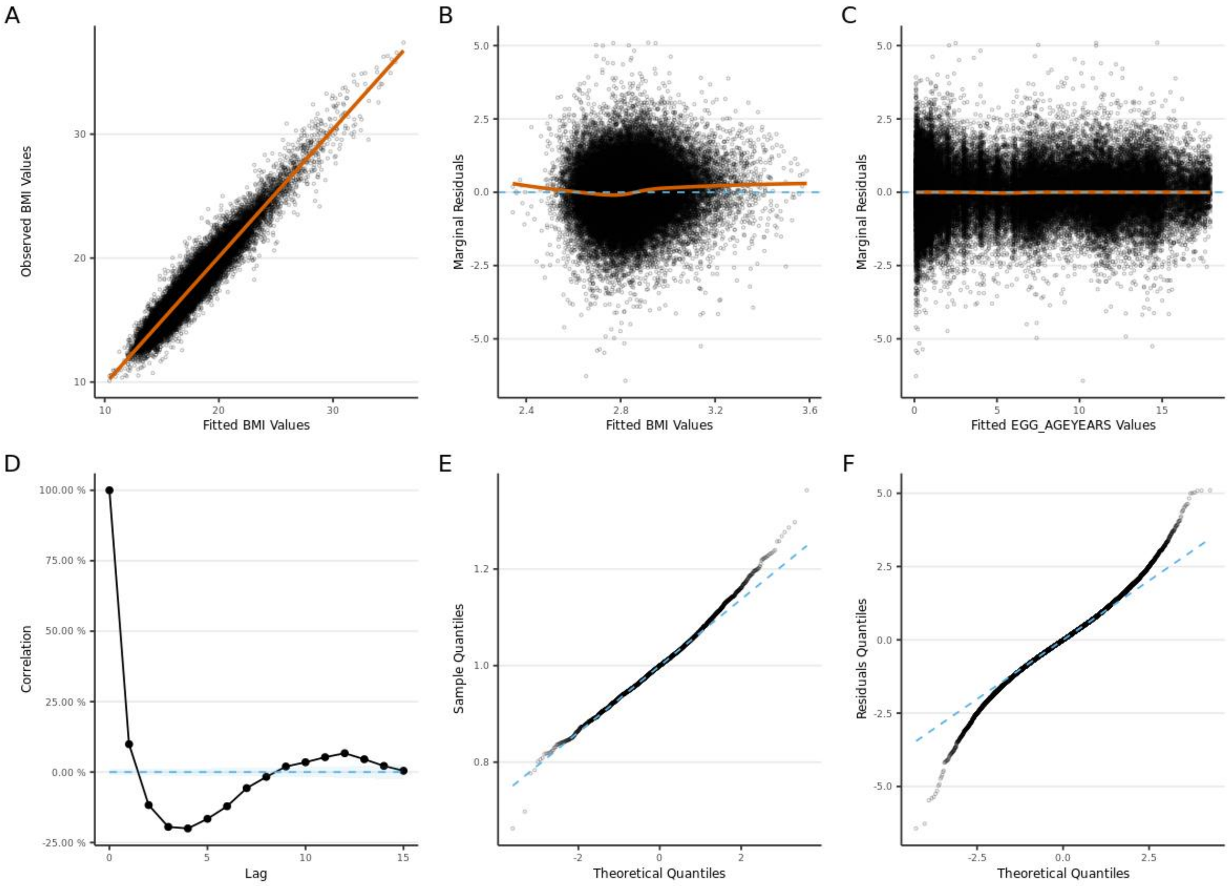
A
Observed BMI Values
Fitted BMI Values
B
Marginal Residuals
Fitted BMI Values
C
Marginal Residuals
Fitted EGG_AGEYEARS Values
D
Correlation
Lag
E
Sample Quantiles
Theoretical Quantiles
F
Residuals Quantiles
Theoretical Quantiles

5: NFBC1986

Male

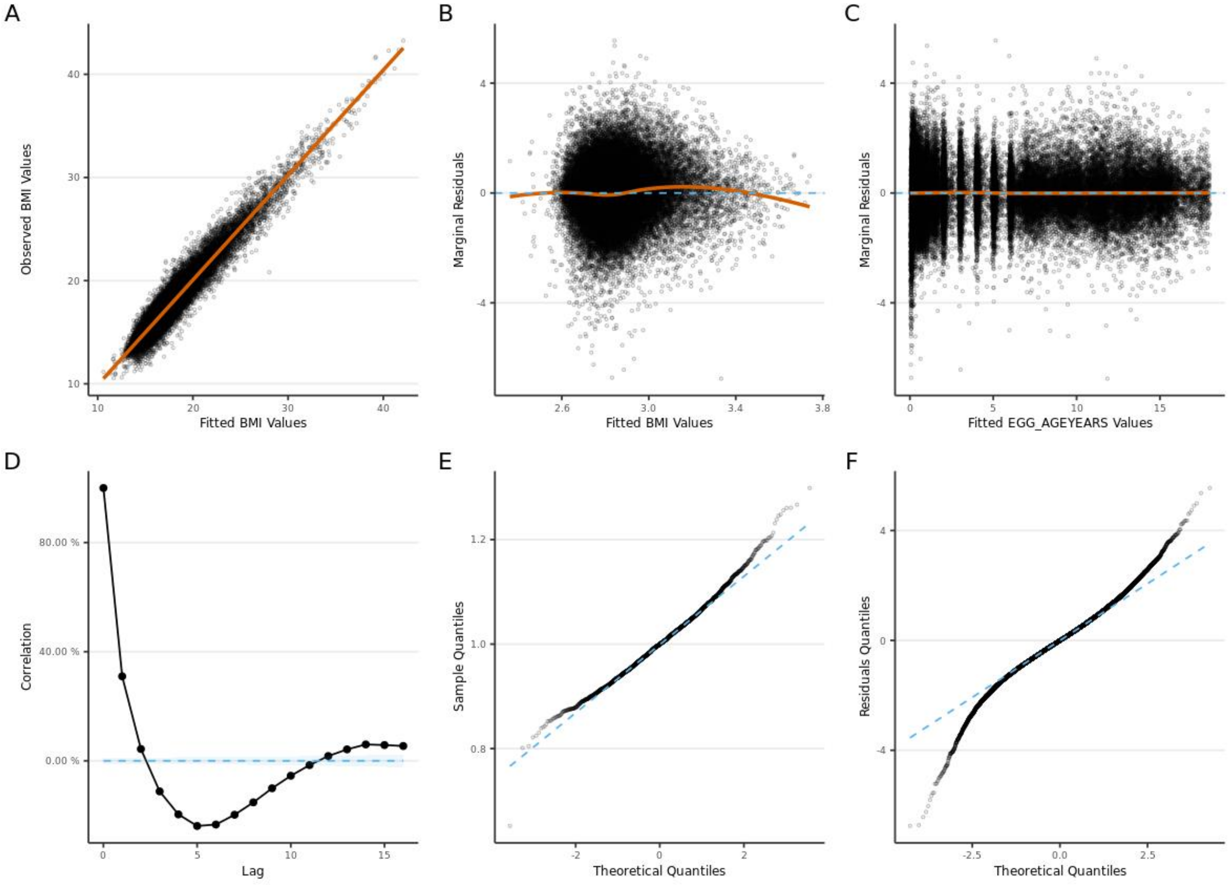

Female

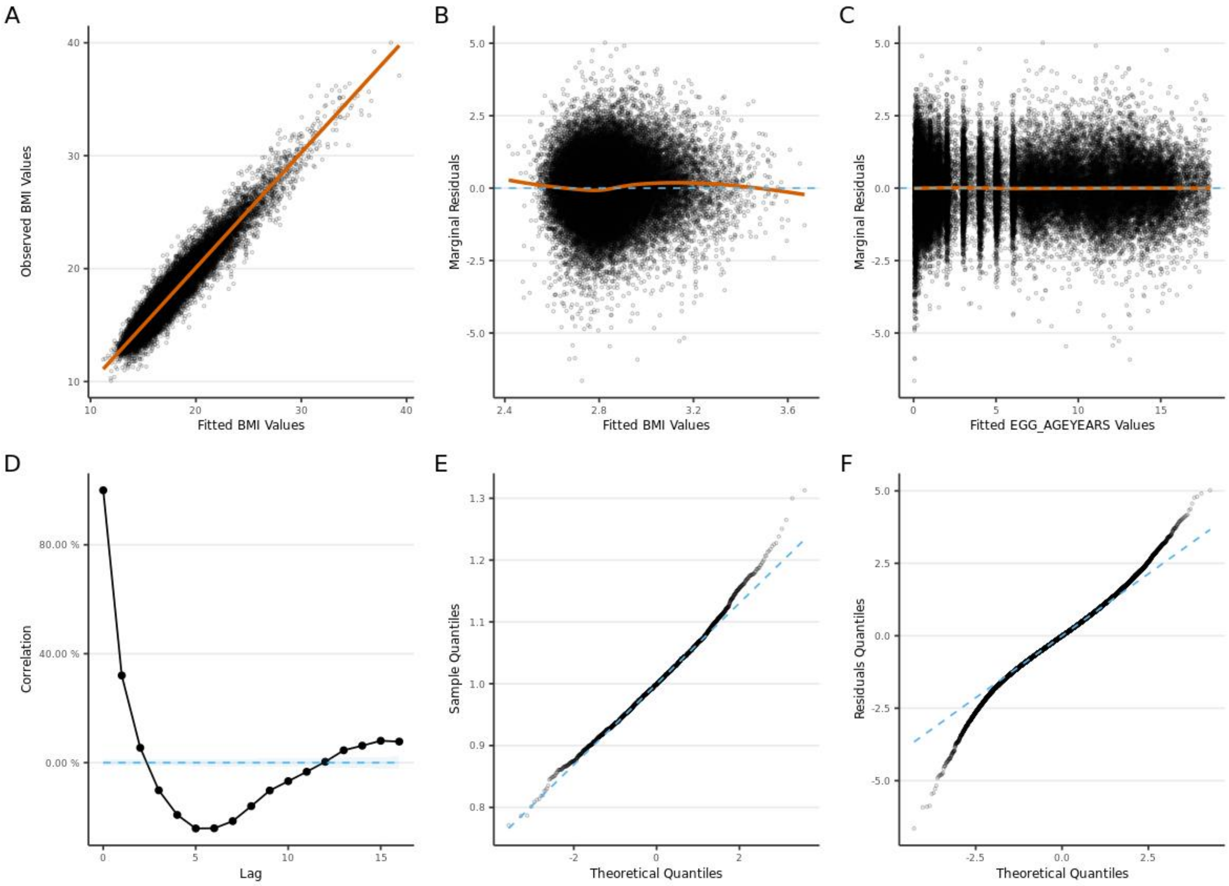

A
B
C
D
E
F
Observed BMI Values
Fitted BMI Values
Marginal Residuals
Fitted BMI Values
Marginal Residuals
Fitted EGG_AGEYEARS Values
Correlation
Lag
Sample Quantiles
Theoretical Quantiles
Residuals Quantiles
Theoretical Quantiles

6: OBE

Male

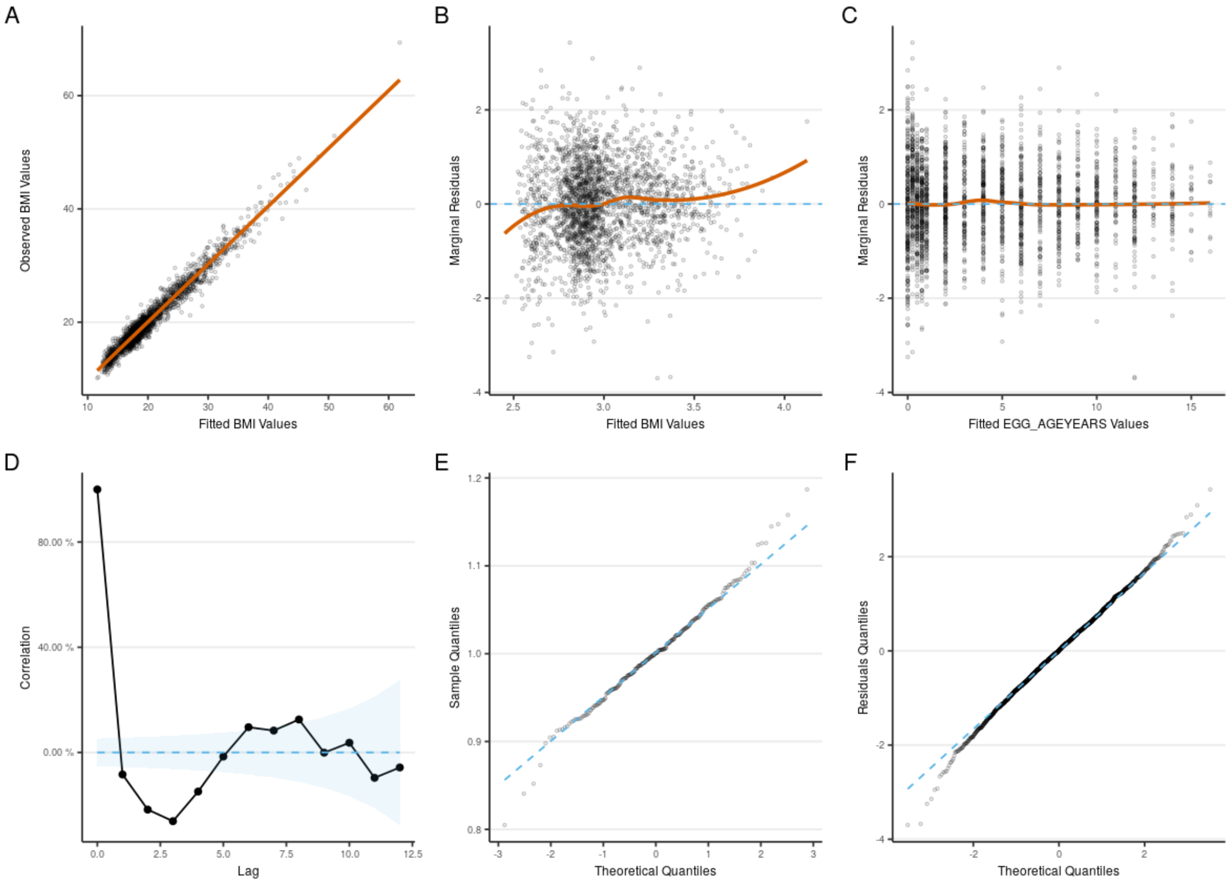
A
B
C
D
E
F
Observed BMI Values
Fitted BMI Values
Marginal Residuals
Fitted EGG_AGEYEARS Values
Correlation
Lag
Sample Quantiles
Theoretical Quantiles
Residuals Quantiles

Female

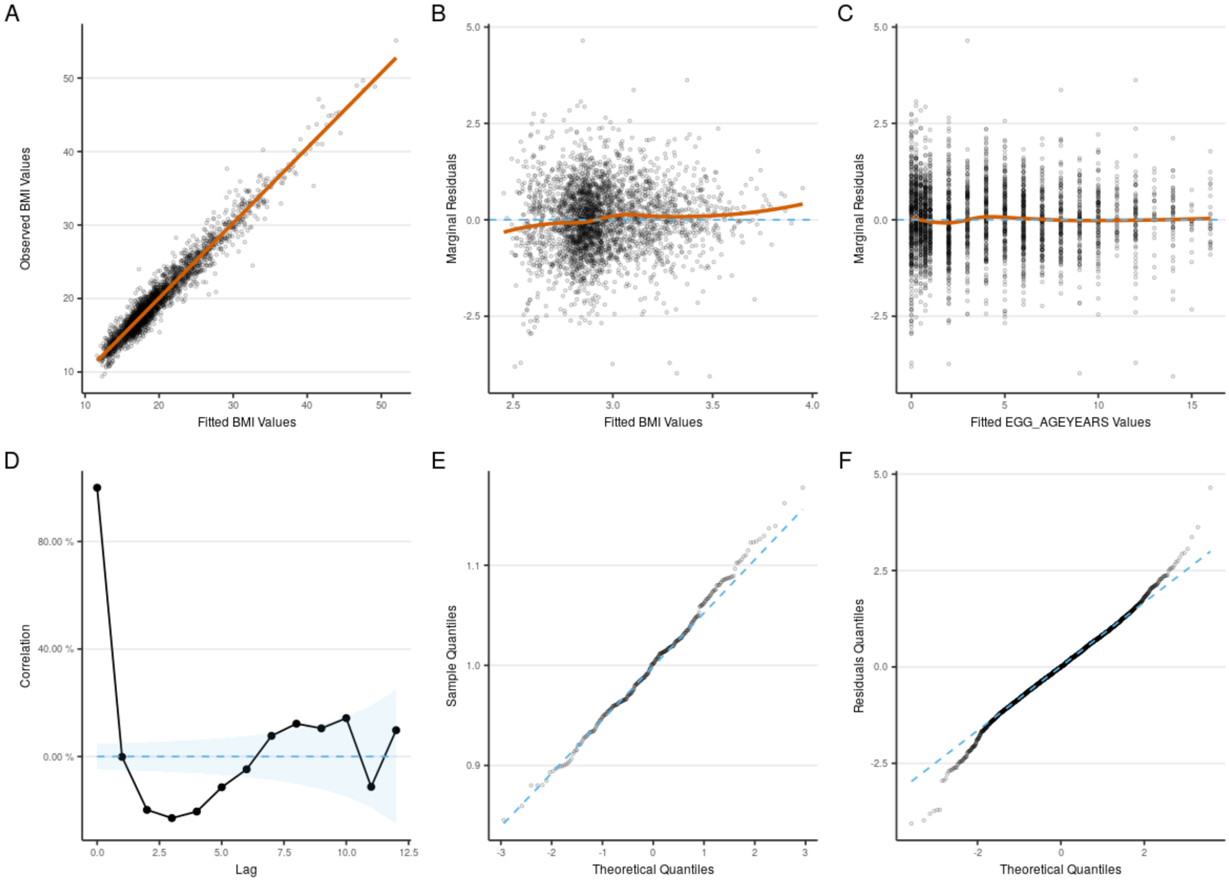
A
B
C
D
E
F
Observed BMI Values
Fitted BMI Values
Marginal Residuals
Fitted BMI Values
Marginal Residuals
Fitted EGG_AGEYEARS Values
Correlation
Lag
Sample Quantiles
Theoretical Quantiles
Residuals Quantiles
Theoretical Quantiles

Supplementary Figure 4: Summary of the association between final BMI (between age 16-18 years) and each of the estimated phenotypes, including a random effects meta-analysis (DerSimonian-Laird estimator).

Within each cohort, both final BMI and the estimated phenotypes were standardized and analyses were adjusted for sex.

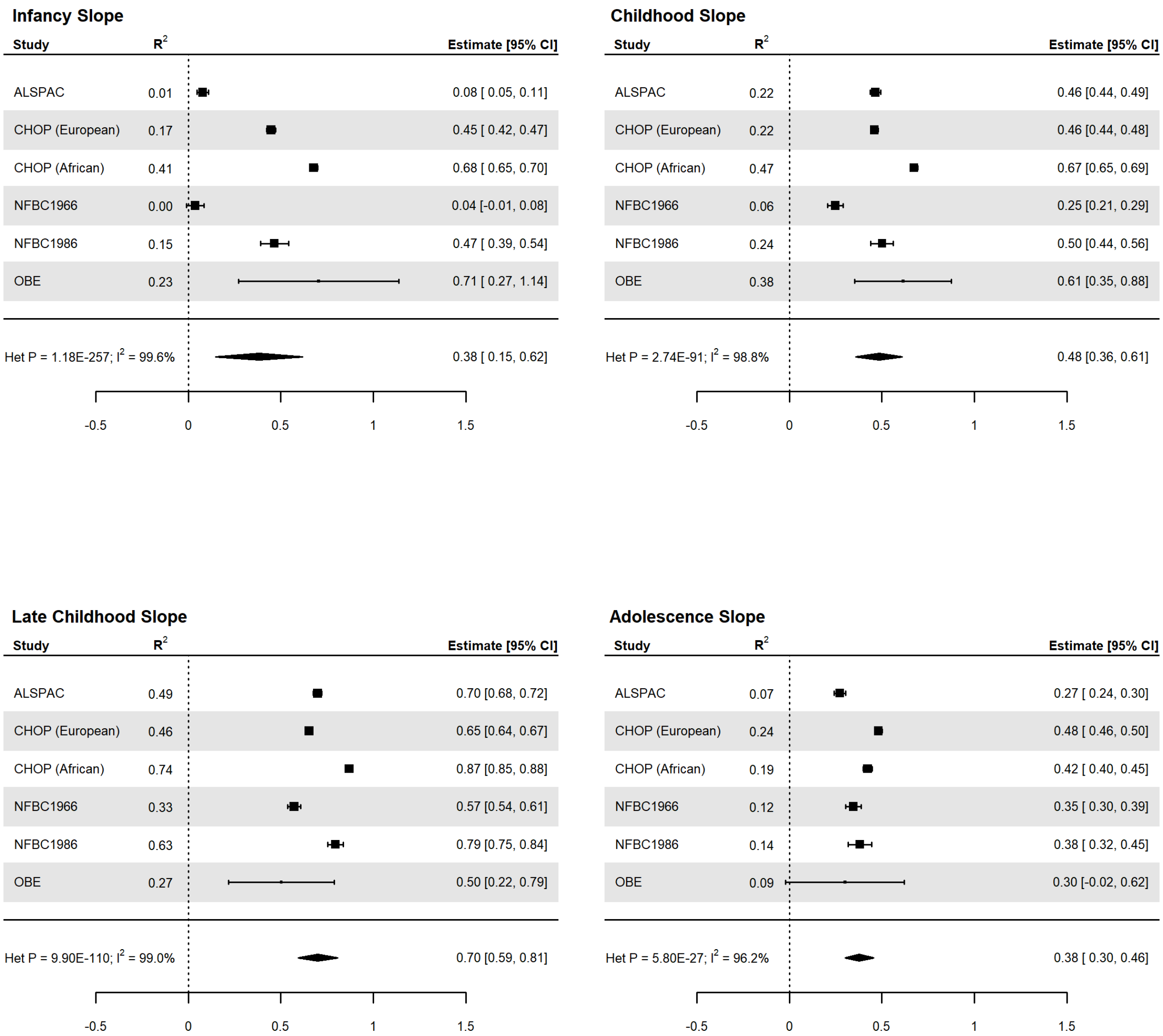

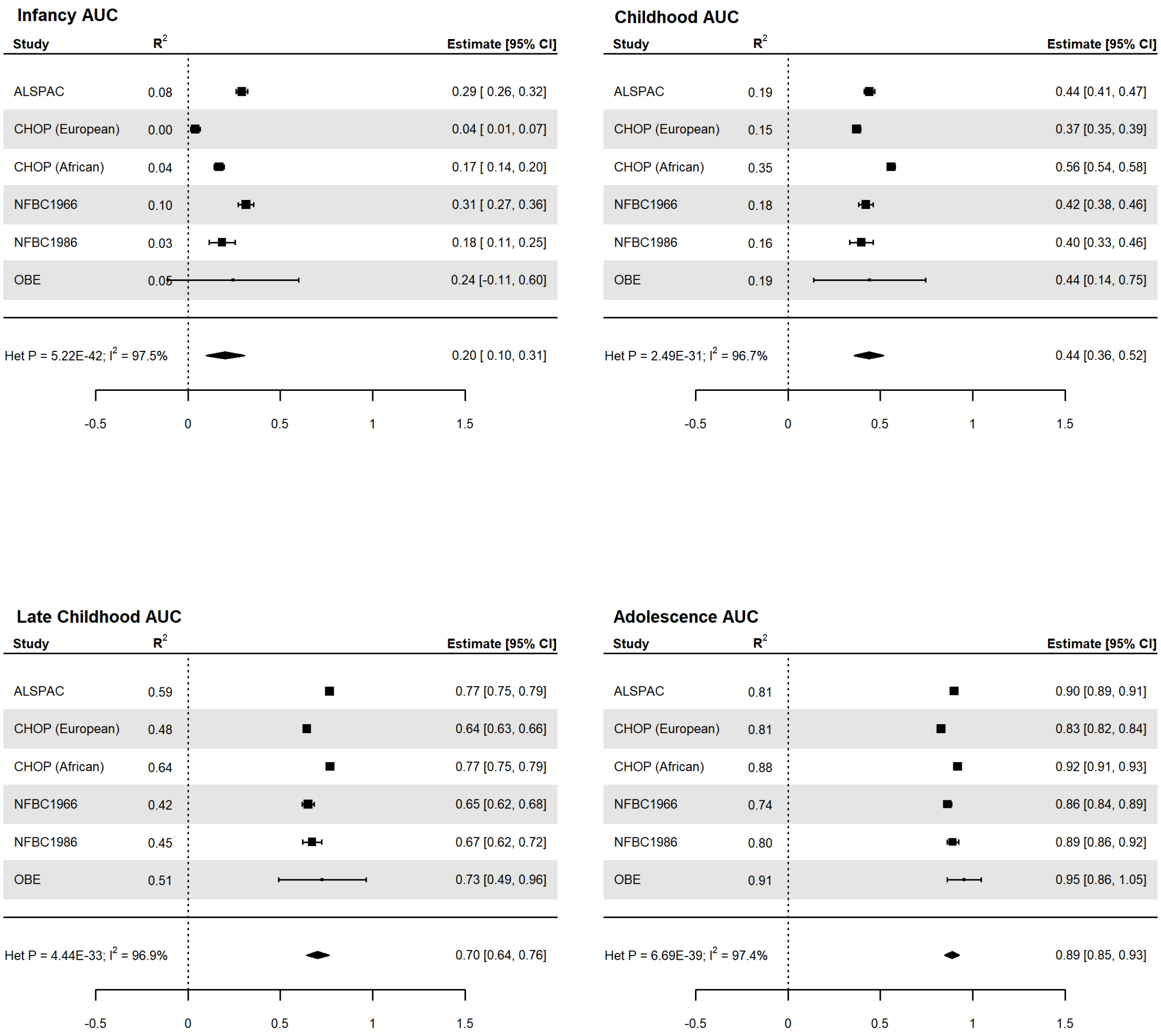
Infancy AUC
Study
R2
Estimate [95% CI]
ALSPAC 0.08 0.29 [ 0.26, 0.32]
CHOP (European) 0.00 0.04 [ 0.01, 0.07]
CHOP (African) 0.04 0.17 [ 0.14, 0.20]
NFBC1966 0.10 0.31 [ 0.27, 0.36]
NFBC1986 0.03 0.18 [ 0.11, 0.25]
OBE 0.05 0.24 [-0.11, 0.60]
Het P = 5.22E-42; I2 = 97.5% 0.20 [ 0.10, 0.31]
-0.5
0
0.5
1
1.5
Childhood AUC
Study
R2
Estimate [95% CI]
ALSPAC 0.19 0.44 [0.41, 0.47]
CHOP (European) 0.15 0.37 [0.35, 0.39]
CHOP (African) 0.35 0.56 [0.54, 0.58]
NFBC1966 0.18 0.42 [0.38, 0.46]
NFBC1986 0.16 0.40 [0.33, 0.46]
OBE 0.19 0.44 [0.14, 0.75]
Het P = 2.49E-31; I2 = 96.7% 0.44 [0.36, 0.52]
-0.5
0
0.5
1
1.5
Late Childhood AUC
Study
R2
Estimate [95% CI]
ALSPAC 0.59 0.77 [0.75, 0.79]
CHOP (European) 0.48 0.64 [0.63, 0.66]
CHOP (African) 0.64 0.77 [0.75, 0.79]
NFBC1966 0.42 0.65 [0.62, 0.68]
NFBC1986 0.45 0.67 [0.62, 0.72]
OBE 0.51 0.73 [0.49, 0.96]
Het P = 4.44E-33; I2 = 96.9% 0.70 [0.64, 0.76]
-0.5
0
0.5
1
1.5
Adolescence AUC
Study
R2
Estimate [95% CI]
ALSPAC 0.81 0.90 [0.89, 0.91]
CHOP (European) 0.81 0.83 [0.82, 0.84]
CHOP (African) 0.88 0.92 [0.91, 0.93]
NFBC1966 0.74 0.86 [0.84, 0.89]
NFBC1986 0.80 0.89 [0.86, 0.92]
OBE 0.91 0.95 [0.86, 1.05]
Het P = 6.69E-39; I2 = 97.4% 0.89 [0.85, 0.93]
-0.5
0
0.5
1
1.5

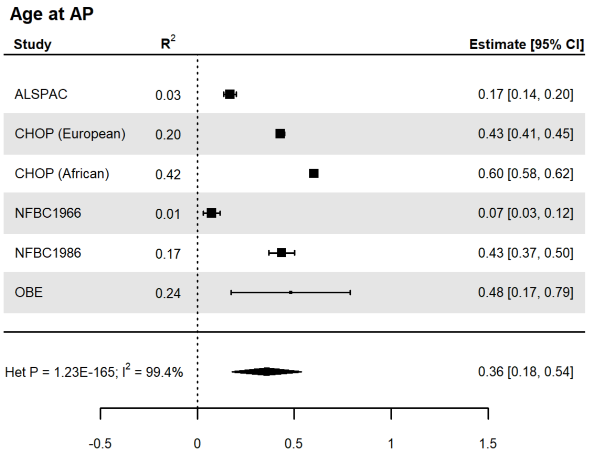
Age at AP
Study
$R^2$
Estimate [95% CI]
ALSPAC 0.03 0.17 [0.14, 0.20]
CHOP (European) 0.20 0.43 [0.41, 0.45]
CHOP (African) 0.42 0.60 [0.58, 0.62]
NFBC1966 0.01 0.07 [0.03, 0.12]
NFBC1986 0.17 0.43 [0.37, 0.50]
OBE 0.24 0.48 [0.17, 0.79]
Het P = 1.23E-165; $I^2$ = 99.4% 0.36 [0.18, 0.54]
-0.5
0
0.5
1
1.5

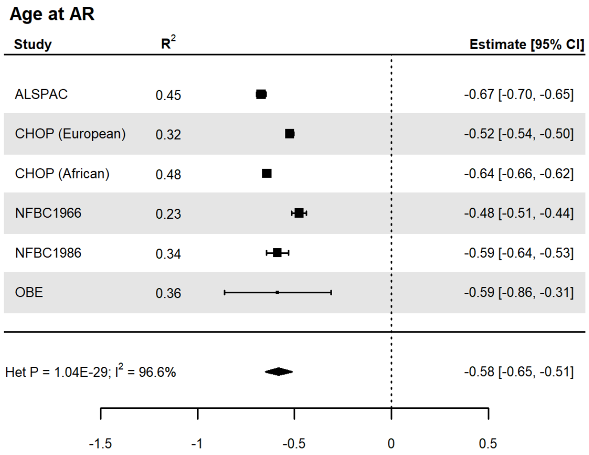
Age at AR
Study
$R^2$
Estimate [95% CI]
ALSPAC 0.45 -0.67 [-0.70, -0.65]
CHOP (European) 0.32 -0.52 [-0.54, -0.50]
CHOP (African) 0.48 -0.64 [-0.66, -0.62]
NFBC1966 0.23 -0.48 [-0.51, -0.44]
NFBC1986 0.34 -0.59 [-0.64, -0.53]
OBE 0.36 -0.59 [-0.86, -0.31]
Het P = 1.04E-29; $I^2$ = 96.6% -0.58 [-0.65, -0.51]
-1.5
-1
-0.5
0
0.5

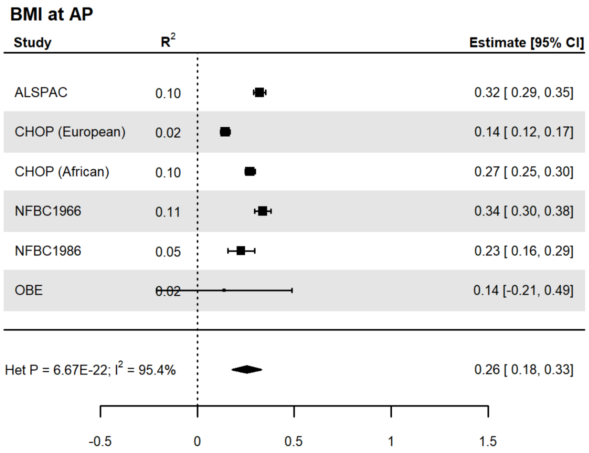
BMI at AP
Study
$R^2$
Estimate [95% CI]
ALSPAC 0.10 0.32 [ 0.29, 0.35]
CHOP (European) 0.02 0.14 [ 0.12, 0.17]
CHOP (African) 0.10 0.27 [ 0.25, 0.30]
NFBC1966 0.11 0.34 [ 0.30, 0.38]
NFBC1986 0.05 0.23 [ 0.16, 0.29]
OBE 0.02 0.14 [-0.21, 0.49]
Het P = 6.67E-22; $I^2$ = 95.4% 0.26 [ 0.18, 0.33]
-0.5
0
0.5
1
1.5

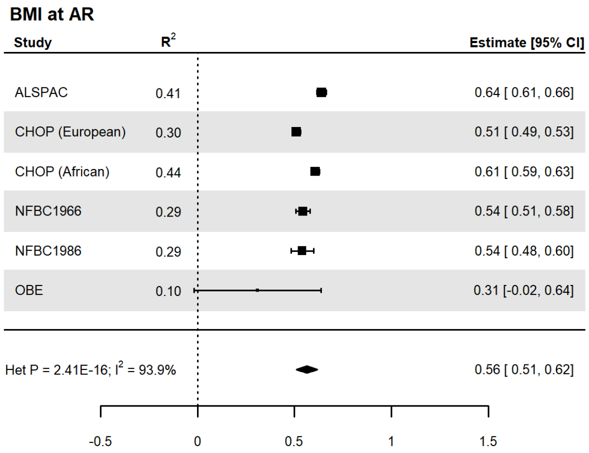
BMI at AR
Study
$R^2$
Estimate [95% CI]
ALSPAC 0.41 0.64 [ 0.61, 0.66]
CHOP (European) 0.30 0.51 [ 0.49, 0.53]
CHOP (African) 0.44 0.61 [ 0.59, 0.63]
NFBC1966 0.29 0.54 [ 0.51, 0.58]
NFBC1986 0.29 0.54 [ 0.48, 0.60]
OBE 0.10 0.31 [-0.02, 0.64]
Het P = 2.41E-16; $I^2$ = 93.9% 0.56 [ 0.51, 0.62]
-0.5
0
0.5
1
1.5

Supplementary Figure 5: Manhattan plots of the meta-analyses without OBE for the phenotypes across infancy (0-0.5 years), early childhood (1.5-3.5 years), late childhood (6.5-10 years) and adolescence (12-17 years).

Loci are labelled with their nearest gene annotated by LocusZoom. The red line corresponds to the genome-wide significance level of $P<5x10^{-8}$

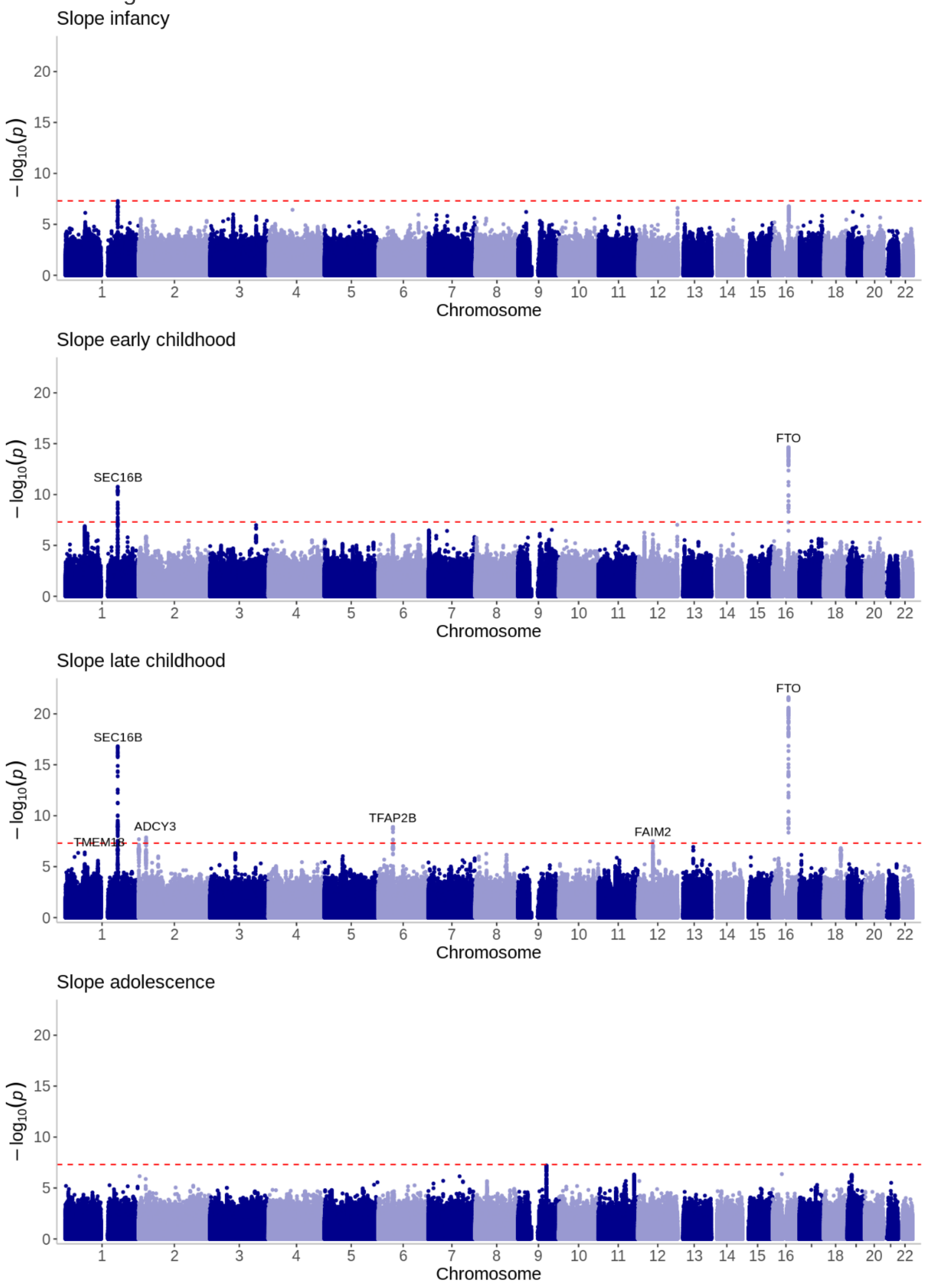

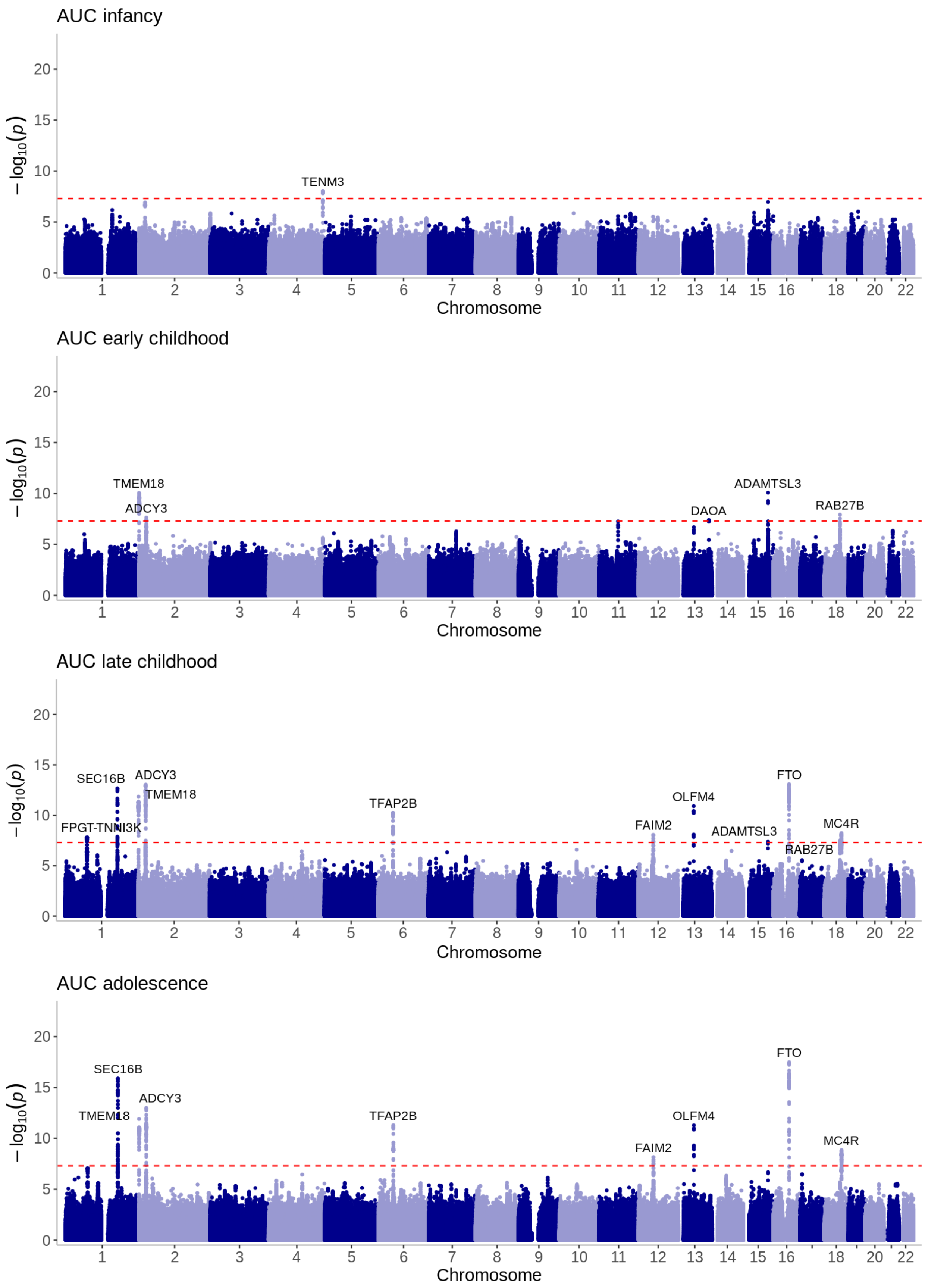
AUC infancy
TENM3
AUC early childhood
TMEM18
ADCY3
DAOA
ADAMTSL3
RAB27B
AUC late childhood
SEC16B
ADCY3
TMEM18
FPGT-TNNI3K
TFAP2B
FAIM2
OLFM4
ADAMTSL3
FTO
RAB27B
MC4R
AUC adolescence
SEC16B
TMEM18
ADCY3
TFAP2B
FAIM2
OLFM4
FTO
MC4R
$-\log_{10}(p)$
Chromosome

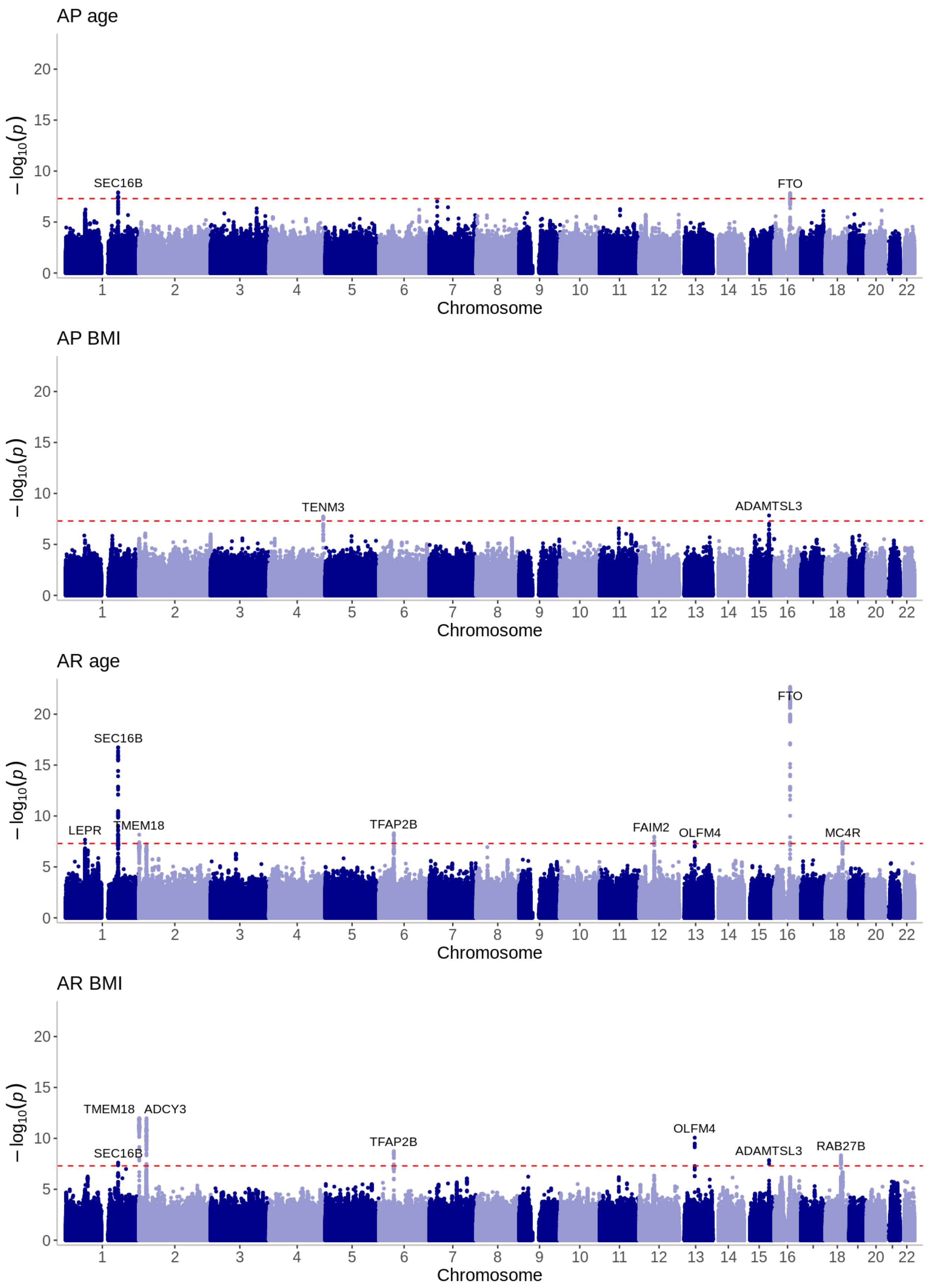
AP age
SEC16B
FTO
AP BMI
TENM3
ADAMTSL3
AR age
SEC16B
FTO
LEPR
TMEM18
TFAP2B
FAIM2
OLFM4
MC4R
AR BMI
TMEM18
ADCY3
SEC16B
OLFM4
TFAP2B
ADAMTSL3
RAB27B
− log10(p)
Chromosome
18
20
22
0
5
10
15
20

Supplementary Figure 6: LocusZoom regional plot of the association between adolescent slope and the FAM120AOS locus

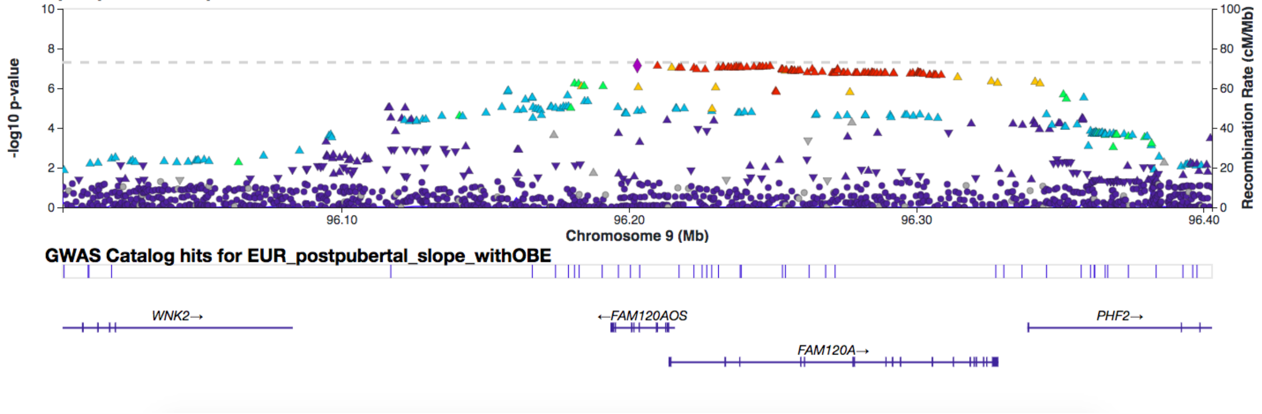

**Cohort description, acknowledgements and funding**

Avon Longitudinal Study of Parents and Children (ALSPAC)

Pregnant women resident in Avon, UK with expected dates of delivery between 1st April 1991 and 31st December 1992 were invited to take part in the study (Boyd et al. 2013; Fraser et al. 2013). The initial number of pregnancies enrolled was 14,541. Of the initial pregnancies, there was a total of 14,676 foetuses, resulting in 14,062 live births and 13,988 children who were alive at 1 year of age. When the oldest children were approximately 7 years of age, an attempt was made to bolster the initial sample with eligible cases who had failed to join the study originally. As a result, when considering variables collected from the age of seven onwards (and potentially abstracted from obstetric notes) there are data available for more than the 14,541 pregnancies mentioned above: The number of new pregnancies not in the initial sample (known as Phase I enrolment) that are currently represented in the released data and reflecting enrolment status at the age of 24 is 906, resulting in an additional 913 children being enrolled (456, 262 and 195 recruited during Phases II, III and IV respectively). The total sample size for analyses using any data collected after the age of seven is therefore 15,447 pregnancies, resulting in 15,658 foetuses. Of these 14,901 children were alive at 1 year of age.

Please note that the study website contains details of all the data that is available through a fully searchable data dictionary and variable search tool:

http://www.bristol.ac.uk/alspac/researchers/our-data/

From birth to approximately five years of age, length and weight measurements were abstracted from health visitor records. Health visits form part of the standard childcare in the UK and data was abstracted from records at approximately six weeks, 10, 21, and 48 months of age. From ages 4 months to 5 years a sub-study (called “Child in Focus”) included a random 10% of the cohort and recorded data across eight research clinics. Length/height and weight were also measured at these clinics. From age 7 onwards, all children were invited to annual research clinics in which height and weight were measured. Furthermore, parent-reported height and weight was also recorded via questionnaires across the time course. BMI was derived from the length/height and weight measures (mean of 9 measures per participant) as weight (kg) divided by the square of height (m). Given the inclusion of measured and self-reported height and weight data, we additionally included an indicator of the source of data as a covariable in the LMMs (clinic/health visit vs. questionnaire). All participants with at least one measure of BMI was included in the study, however, we excluded those who were part of a multiple birth (i.e. twins, triplets etc. n=524).

Ethical approval for the study was obtained from the ALSPAC Ethics and Law Committee and the Local Research Ethics Committees. Informed consent for the use of data collected via questionnaires and clinics was obtained from participants following the recommendations of the ALSPAC Ethics and Law Committee at the time.

We are extremely grateful to all the families who took part in this study, the midwives for their help in recruiting them, and the whole ALSPAC team, which includes interviewers, computer and laboratory technicians, clerical workers, research scientists, volunteers, managers, receptionists and nurses.

The UK Medical Research Council and Wellcome (Grant ref: 217065/Z/19/Z) and the University of Bristol provide core support for ALSPAC. A comprehensive list of grants funding is available on the ALSPAC website (http://www.bristol.ac.uk/alspac/external/documents/grant-acknowledgements.pdf). Genome-wide genotyping data was generated by Sample Logistics and Genotyping Facilities at Wellcome Sanger Institute and LabCorp (Laboratory Corporation of America) using support from 23andMe. This publication is the work of the authors and Kimberley Burrows will serve as guarantors for the contents of this paper.

For the purpose of Open Access, the author has applied a CC BY public copyright license to any Author Accepted Manuscript version arising from this submission.

Northern Finland Birth Cohorts born in 1966 (NFBC1966)
The Northern Finland Birth Cohort study 1966 (NFBC1966) includes 12,058 live born individuals, of European descent, with expected dates of birth during 1966 in the two northernmost provinces of Finland, Oulu and Lapland (1966 University of Oulu. 1966). The data on all cohort members were prospectively collected since pregnancy and supplemented at the ages of 1, 14, 31 and 46 years (Nordström et al. 2021). Height and weight growth measurements were obtained from communal child health clinics (Sovio et al. 2009). All those living in northern Finland or in the capital area were invited to a clinical examination and blood sampling at age 31 years. DNA was extracted from 5753 individuals and Illumina's HumanCNV370-Duo DNA Analysis BeadChip was used to obtain genome-wide genetic data (Sabatti et al. 2009).

Prior to growth trajectory modelling, we excluded participants born pre-term (n=765) and non-singletons (n=197). We also identified the sibling pairs of this cohort that were born at different times, and one participant per sibling pair was excluded (n=17).

Approval for the studies was granted by the Northern Ostrobothnia Hospital District Ethical Committee 94/2011 (12.12.2011), Finland in accordance with the declaration of Helsinki. Mothers gave their informed consent in the beginning of the NFBC1966 data collection. Written informed consent has been obtained from the cohort participants in the 31- and 46-year data collections.

We thank all cohort members and researchers who participated in the study. We also wish to acknowledge the work of the NFBC project center. NFBC1966 31-year follow-up received financial support from University of Oulu Grant no. 65354, Oulu University Hospital Grant no. 2/97, 8/97, Ministry of Health and Social Affairs Grant no. 23/251/97, 160/97, 190/97, National Institute for Health and Welfare, Helsinki Grant no. 54121, Regional Institute of Occupational Health, Oulu, Finland Grant no. 50621, 54231. NFBC1966 46yr follow-up received financial support from University of Oulu Grant no. 24000692, Oulu University Hospital Grant no. 24301140, ERDF European Regional Development Fund Grant no. 539/2010 A31592.

Northern Finland Birth Cohorts born in 1986 (NFBC1986)
The Northern Finland Birth Cohort 1986 (NFBC1986) includes 9,432 live born children with expected dates of birth between 1st July 1985 and 30th June 1986 in the two northernmost provinces of Finland, Oulu and Lapland (Järvelin, Hartikainen-Sorri, and Rantakallio 1993; Järvelin et al. 1997). The cohort has been followed up since early pregnancy until adulthood (1986 University of Oulu. 1986). Growth measurements were obtained from communal child health clinics. All those alive with known address were invited to a clinical examination at the age of 15 to 16 years. At this age, blood samples were drawn and DNA was extracted for 6,266 subjects.

Prior to growth trajectory modelling, we excluded pre-term babies (n=496) as well as twins and triplets (n=132). From the remaining sample, we further identified one sibling pair that was born at different times, and the child with less height and weight measurements was excluded. Finally, one child with extremely fast BMI growth from early on was excluded from the sample as the trajectory was not comparable to other children.

Approval for the studies was granted by the Northern Ostrobothnia Hospital District Ethical Committee 108/2017 (15.1.2018), Finland in accordance with the declaration of Helsinki. Participants provided written informed consent in data collections carried out when they were aged 15 to 16 and 33 to 35 years.

We thank all cohort members and researchers who have participated in the study. We also wish to acknowledge the work of the NFBC project center. The cohort received financial support from the following grants EU QLG1-CT-2000-01643 (EUROBLCS) Grant no. E51560, NorFA Grant no. 731, 20056, 30167, USA / NIH 2000 G DF682 Grant no. 50945.

CHOP
The CHOP cohort is a random sampling of the population of children who come for care at the Children's Hospital of Philadelphia starting in 2006 and continuing presently (Connolly et al. 2020). If patients choose to enroll in the study, their fully de identified and encrypted electronic medical records are available for study. Heights and weights used to calculate BMI in this study were obtained from electronic medical records. Self-reported ancestry was used to define African Americans and Europeans Americans. The CHOP study was genotyped at the Center for Applied Genomics at the Children's Hospital of Philadelphia. The Research Ethics Board of CHOP approved the study, and written informed consent was obtained from all subjects.

The authors thank the network of primary care clinicians and the patients and families for their contribution to this project and to clinical research facilitated by the Pediatric Research Consortium [PeRC]-The Children's Hospital of Philadelphia. R. Chiavacci, E. Dabaghyan, A. [Hope] Thomas, K. Harden, A. Hill, C. Johnson-Honesty, C. Drummond, S. Harrison, F. Salley, C. Gibbons, K. Lilliston, C. Kim, E. Frackelton, F. Mentch, G. Otieno, K. Thomas, C. Hou, K. Thomas and M.L. Garris provided expert assistance with genotyping and/or data collection and management. The authors would also like to thank S. Kristinsson, L.A. Hermannsson and A. Krisbjörnsson of Raförninn ehf for extensive software design and contributions. This research was financially supported by an Institute Development Award from the Children's Hospital of Philadelphia, a Research Development Award from the Cotswold Foundation, the Daniel B. Burke Endowed Chair for Diabetes Research, the Children's Hospital of Philadelphia Endowed Chair in Genomic Research and NIH grant R01 HD056465.

OBE
The OBésité de l'Enfant (OBE) cohort includes children with early-onset, familial obesity, who were born in France. Most of them were recruited by the CNRS 8199 unit in 1998. A small part of

these children were patients of Toulouse Children's Hospital. Height and weight measurements were taken by the participant's GP or pediatrician. The study protocols were approved by local ethics committees. Oral assent from children was obtained and parents (or legal guardians) signed an informed consent form.
The authors would like to thank the French National Research Agency (Agence Nationale de la Recherche [ANR]-10-LABX-46 [European Genomics Institute for Diabetes] and ANR-10-EQPX-07-01 [Lille Integrated Genomics Advanced Network for personalized medicine] to A.B. and P.F.), European Research Council (OpiO 101043671 to A.B.), the European Union's Horizon Europe Research and Innovation Programme (OBELISK grant agreement 101080465 to A.B. and P.F.), and the National Center for Precision Diabetic Medicine (PreciDIAB to A.B. and P.F), which is jointly supported by the French National Agency for Research (ANR-18-IBHU-0001), European Regional Development Fund, Hauts-de-France Regional Council, and the European Metropolis of Lille. The authors also thank the France Génomique consortium (ANR-10-INBS-009).

**Linear mixed modelling**

A simple approach is to model a repeated continuous outcome (e.g. BMI) as a function of time within a linear mixed-effects model (LMM). LMM has an advantage over other methods, such as repeated measures ANOVA, by allowing missing data within individuals, and unbalanced study designs where either the number of measures or the timing of the measurements differs between individuals, as well as fitting and testing covariance structures (Laird and Ware 1982).

A basic random-slope LMM with a single continuous outcome such as BMI as a linear function of time can be written as:

$$Y_{it} = \beta_0 + \beta_1 X_{it} + \upsilon_{0i} + \upsilon_{1i} X_{it} + \varepsilon_{it}$$

Where $Y_{it}$ is a single outcome measured for individual *i* at time *t* and is assumed to be independent between individuals. The fixed coefficients, $\beta_0$ and $\beta_1$ represent the average intercept and slope respectively. The random coefficients represent the deviation from the average intercept ($\upsilon_{0i}$) and slope ($\upsilon_{1i}$) for individual *i*. The random effects are assumed to follow a bivariate normal distribution with mean zero and covariance matrix $\Omega_\upsilon$. Residual errors $\varepsilon_{it}$ are assumed to be independently identically normally (i.i.d) distributed with variance $\sigma_{e0}^2$.

The LMM model above assumes a linear change in the continuous outcome with increasing time. However, growth is nonlinear for many biological characteristics, such as BMI across early life. BMI throughout infancy, childhood, and adolescence follows a complex pattern of change including the appearance of the adiposity peak during infancy and then a rebound during childhood (see Figure 1 in the main text). This nonlinear change can be incorporated into LMM models through the addition polynomial functions of time. A quadratic function for instance can induce a curve with a single turn, while a cubic function can involve up to two turns. More simple polynomials may not adequately describe the turning points in the underlying data, while more complex polynomials can produce artefactual turns in the curve and can particularly fit poorly at extremes where data is more sparse. A more flexible approach to model complex curves such as BMI is through using spline functions, joined at knot points dispersed across time. Smooth functional polynomials (usually of low order) are chosen to fit the measures between consecutive knots (Perperoglou et al. 2019).

**Example process and output from the *EGGLA* BMI modelling framework**

While the framework described below is specific to the current study exploring the GWAS of longitudinal BMI trajectories and estimated phenotypes, the framework can serve as an exemplar for other consortium efforts to analyse other non-linear longitudinal traits.

To undertake LMM analysis of BMI, analysts are required to load in longitudinal data in long format with the following mandatory columns: individual ID, sex, height measurement (cm), weight measurement (kg), and age at measurement (days or years). Additional cohort specific covariates may also be included.

A comprehensive protocol has been written to allow the user to set up the environment and work through the analysis pipeline. This is available at: https://m.canouil.dev/eggla/articles/eggla.html. The online protocol contains a series of vignettes containing the code used to perform the model diagnostics framework (https://m.canouil.dev/eggla/articles/models-diagnostics.html), a plot to aid in model selection (https://m.canouil.dev/eggla/articles/model-selection.html), analyses and plots specific to the chosen model from the diagnostics framework (in this case specific to the cubic spline model) (https://m.canouil.dev/eggla/articles/run-cubic-splines.html), and finally characterisation of the AP and AR (https://m.canouil.dev/eggla/articles/adiposity-peak-rebound.html). See each vignette for examples of output using simulated data.

The chosen best model selected from the output of the vignettes above was taken forward for model refinement (see methods of main paper). Finally, two *EGGLA* R package (Canouil et al. 2024) functions; run_eggla_lmm() and run_eggla_gwas() were used to perform the LMM and GWAS using the final chosen model (cubic spline function in the fixed effects with cubic slope function random effects).

The *EGGLA* (Canouil et al. 2024) function run_eggla_lmm() is a wrapper used to perform the following analysis stages: 1) cleaning and formatting of BMI data using the *growthcleanr* R package for longitudinal data cleaning, 2) deriving BMI from cleaned height and weight data and formatting of the dataframe for analysis, 3) run the LMM for males and females separately, 4) plotting of model residuals, 5) deriving slopes and AUCs for defined linear time intervals of the curve, 6) predict BMI and derive age and BMI at AP and AR, 7) detection of outlier individuals for each of the estimated phenotypes and output of correlations between latent phenotypes, 8) formatting the derived phenotypes into a dataset ready for GWAS. The run_eggla_lmm() function allows for further refinement of models where applicable including: addition of covariates, choice of knot points and placement, choice of the linear time periods for derivation of slopes and AUCs, inclusion of correlation structure and complexity of random effects. After specifying the parameters of the chosen model (knot points, time intervals, covariates, number of iterations, and parameters for outliers) and running the model using the *run_eggla_lmm()* R function, several outputs are provided to aid the analyst in making a final checks; these outputs include plots of residuals, the model coefficients, the model call, a table of pairwise correlations between all derived phenotypes and a list of outliers identified for each of the latent variables. This allows the analyst to make decisions about model fit, outlier inclusion or exclusion and inform downstream analysis based on the extent of the correlation between the derived phenotypes. Finally, individual level derived phenotypes for each specified time-period are output as comma-separated files. For instance a .csv file will contain all individuals (males and females have a separate .csv file) and their slopes for infancy, childhood, late childhood, and adolescence. The csv files can then be called by the *run_eggla_gwas()* R function or the sex stratified files can be combined and included in the analysts preferred software for GWAS.

The run_eggla_gwas() function performs GWAS analysis that can be implemented by any cohort with unrelated individuals (any complex data structures, such as related family structure, will need to use alternative GWAS software), where genetic dosage data is available in VCF file format. The function analyses VCF files through software packages BFCtools (v.≥1.16) (https://github.com/samtools/BCFtools) and PLINK2 (v.≥2.0) (https://www.cog-genomics.org/plink/2.0) and therefore requires access to these packages. The following actions are performed using the run_eggla_gwas() function: 1) format input data and perform checks for ID matching, and removing outliers on a complete case basis if required, 2) format VCFs through BCFtools and annotate if requested, 3) perform GWAS using PLINK2, 4) extract information from VCF INFO field if requested, and 5) allows for further options including:  annotation files to add rsID and gene symbols
